# Cash Transfer Programs and HIV-Related Outcomes: an Analysis of 42 Countries from 1996 to 2019

**DOI:** 10.1101/2021.12.16.21267921

**Authors:** Aaron Richterman, Harsha Thirumurthy

## Abstract

**Background:** Many low- and middle-income countries have introduced cash transfer programs as part of their poverty reduction and social protection strategies. These programs have the potential to overcome various drivers of HIV risk behaviors and usage of HIV services, but their overall effects on a broad range of HIV-related outcomes remains unknown.

**Methods:** We used publicly reported data to determine whether low- and middle-income countries with HIV prevalence >1% and baseline annual incidence >1/1000 had conditional or unconditional cash transfer programs that covered >5% of the impoverished population, and the year in which those programs began and ended. We obtained country- and individual-level data on HIV-related outcomes from UNAIDS and population-representative household surveys, focusing on the period between 1996 and 2019. We conducted difference-in-differences analyses with country and year fixed effects to evaluate the effects of cash transfer programs on country- and individual-level HIV-related outcomes.

**Findings:** Forty-two countries across three continents were included. Among these, 21 were in the intervention group, having implemented cash program(s) with impoverished population coverage greater than 5% during the study period. Cash transfer programs were associated with lower probability of reporting sexually transmitted infection within the last 12 months among females (odds ratio [OR] 0.67, 95% confidence interval [CI] 0.50-0.91) and higher probability of an HIV test within the last 12 months among females (OR 2.61, 95% CI 1.15-5.88) and males (OR 3.19, 95% CI 2.45-4.15). For country-level outcomes, cash transfer programs were associated with a reduction in new HIV infections (incidence rate ratio [IRR] 0.94, 95% CI 0.89-0.99), but not with the proportion of people with HIV receiving antiretroviral therapy (5.0%, 95% CI -0.2-10.1) or AIDS-related deaths (IRR 0.99, 95% CI 0.95-1.03), though temporal analyses showed delayed improvements in both antiretroviral coverage and deaths.

**Interpretations:** Cash transfer programs, which are being expanded in the context of the COVID-19 pandemic, have the potential to promote ongoing efforts to end HIV as a public health threat. Alongside the already existing focus on expanding biomedical services, these anti-poverty programs can play a greater role in achieving global targets for HIV prevention and treatment.

**Funding:** None

## Introduction

HIV continues to be a major global public health threat, causing an estimated 1.7 million new infections and 690,000 AIDS-related deaths in 2019.^1^ The Joint United Nations Programme on HIV/AIDS (UNAIDS) Fast Track plan set the goal in 2014 of reducing annual infections to 200,000 and AIDS-related deaths by 90% by 2030.^2^ In addition to the rapid scaling up of clinical services, the Fast Track plan also emphasizes the importance of expanding social protection for achieving these objectives.^3^ Social protection is thought to be important because of the well-documented relationship between poverty or economic inequality and risk factors for HIV transmission (e.g., transactional sex among adolescent girls and young women, earlier age at sexual debut, lower use of HIV services, and worse antiretroviral adherence) and HIV-related morbidity and mortality.^4-12^

Over the past two decades, many low- and middle-income countries (LMICs) have introduced cash transfer programs as central components of their poverty reduction and social protection strategies. These programs, which range from conditional cash transfer (CCT) programs that are common in Latin America to unconditional cash transfer (UCT) programs that are common in sub-Saharan Africa, exist in over 100 LMICs, and many countries have expanded or introduced new programs during the COVID-19 pandemic. A growing evidence base suggests that cash transfer programs reduce poverty, foster economic autonomy, raise school attendance for children, improve empowerment for women, and increase health service use, among other benefits.^13^ Conceptually, cash transfer programs may improve outcomes by increasing income and addressing economic barriers as well as by alleviating the psychological impacts of poverty on mental bandwidth and decision-making.^14-19^

Despite the existence of cash transfer programs in many countries with generalized HIV epidemics and the large number of evaluations of these programs, relatively few studies have examined their effects on HIV-related outcomes at the individual or population level. Several studies of predominantly smaller-scale cash transfer interventions have examined the direct effects on beneficiaries and shown mixed but generally favorable changes in HIV-related outcomes. For HIV prevention, a few randomized controlled trials of cash transfers have focused on adolescent girls and young women. In Malawi, a cash transfer intervention for schooling reduced HIV prevalence among schoolgirls.^20^ In South Africa, conditional cash transfers for schooling had no effect on HIV incidence among adolescent girls and young women, although the control group in this study received cash transfers and school attendance was high in both study groups.^21^ No randomized controlled trials have studied the impact of unconditional cash transfer programs, which are more commonly used by governments in sub-Saharan Africa, on HIV incidence. Non-experimental impact evaluations of the Kenyan government’s cash transfer program for caregivers of orphans and vulnerable children and another of the Malawi government’s household cash transfer program found delays in sexual debut.^22,23^ Finally, a much larger literature has examined the effects of financial and non-financial incentives that are tailored to specific HIV-related behaviors, but the incentive amounts are typically much smaller than the size of cash transfers typically administered in LMICs. The typical magnitude of these differences can be illustrated in Tanzania, where financial incentives conditional on HIV clinic attendance were studied for six months in 2018 with a maximum total transfer of about 2% of Gross Domestic Product (GDP) per capita, versus the national Productive Social Safety Net Programme, which provides about 12% of GDP per capita annually to poor households.^24,25^

Studies of incentives have had mixed results, as some have demonstrated improvements in HIV testing uptake,^26-29^ retention in care,^25,30,31^ adherence to antiretroviral therapy,^31-33^ and virologic suppression,^25,34^ while others have not.^30,35-37^ An evaluation of large-scale cash transfer programs using cross population-level data from many different countries remains an important gap in the literature. An advantage of using population-level data when evaluating cash transfer programs (rather than using data from cash transfer beneficiaries alone) is that it will be possible to detect spillover effects of cash transfers, especially if the transfers influence health behaviors that affect the risk of HIV acquisition and transmission.

We hypothesized that larger, more generalized cash transfer programs might improve both population and individual HIV-related outcomes. While national cash transfer programs in sub-Saharan Africa are more commonly unconditional and less HIV-specific than those considered in the studies described above, benefits may still be seen because of their more expansive reach and spillover effects stemming from reduced HIV transmission. These national cash transfer programs also tend to persist over time, compared to financial incentive or cash transfer studies in which interventions have tended to be time limited.

However, few studies have evaluated the broader effects of large-scale cash transfer programs, a policy-relevant topic given the burden of HIV and growing reliance on cash transfer programs. To address this unanswered question, we conducted a difference-in-differences analysis evaluating the effects of cash transfer programs on country- and individual-level outcomes in 42 countries with generalized HIV epidemics from 1996 to 2019.

## Methods

We included all countries with HIV incidence greater than 1 per 1000 persons in 1996 and HIV prevalence greater than 1% in at least one year between 1996 and 2019,^1^ a period when many countries introduced cash transfer programs.

### Data

We identified all major cash transfer programs within included countries. We manually searched a variety of sources, detailed in the Supplementary Appendix, to identify the programs and determine the year in which they were introduced, target population, whether they were conditional or unconditional, amount of transfer, and the most recently available number of beneficiaries. For each cash transfer program, we estimated the most recent impoverished population coverage by dividing the total number of beneficiaries by the number of people living below the international poverty line (Supplementary Appendix).

For individual-level data on HIV outcomes, we used the Demographic and Health Surveys (DHS), which are nationally representative cross-sectional household surveys conducted every 5 years in many LMICs (Supplementary Appendix). Information was obtained for household and individual characteristics for all female household members of reproductive age (15-49 years) and a subset of males of reproductive age (typically 15-49, 54, or 59 years). We also used AIDS Indicator Surveys (AIS), which are similar household surveys focused on HIV knowledge, attitudes, behavior, and prevalence. We used DHS or AIS data from any country that met eligibility criteria and any year between 1996 and 2019.

For country-level HIV statistics, we relied on UNAIDS annual estimates that are generated with modeling techniques based on representative population-based surveys and surveillance studies.^1^ We obtained country and year UNAIDS estimates for the number of new HIV infections, the number of AIDS-related deaths, and proportion of people with HIV receiving antiretroviral therapy.

We obtained additional time-varying covariates for each country and year that were likely to be associated with changes in cash transfer programs and HIV outcomes: Gross Domestic Product (GDP) per capita,^38^ President’s Emergency Plan for AIDS Relief (PEPFAR) funding budgeted per capita,^39^ The Global Fund to Fight AIDS, Tuberculosis and Malaria disbursements for HIV-related programs per capita,^40^ and six Worldwide Governance Indicators from The World Bank that are composite indicators based on 30 data sources: Voice and Accountability, Political Stability and Absence of Violence, Government Effectiveness, Regulatory Quality, Rule of Law, and Control of Corruption.^38^

### Statistical Analysis

We performed difference-in-differences analyses using multivariable regression models to compare trends in HIV-related outcomes in countries with cash transfer programs to those in the same countries prior to cash transfer program introduction and to those in comparison countries without cash transfer programs. Our analysis was developed based on a proposed causal framework linking cash transfer programs to HIV-related outcomes, mediated through an effect on poverty (Figure 1). Our primary explanatory variable of interest was a binary variable indicating presence in a given year of a cash transfer program (or combination of programs) for which the number of beneficiaries exceeded 5% of the population living below the poverty line (Supplementary Appendix).^38^ Our choice of 5% impoverished population coverage as the threshold for our intervention group was subjective but chosen empirically as the smallest likely coverage with which we might expect to see population effects.

**Figure 1.**
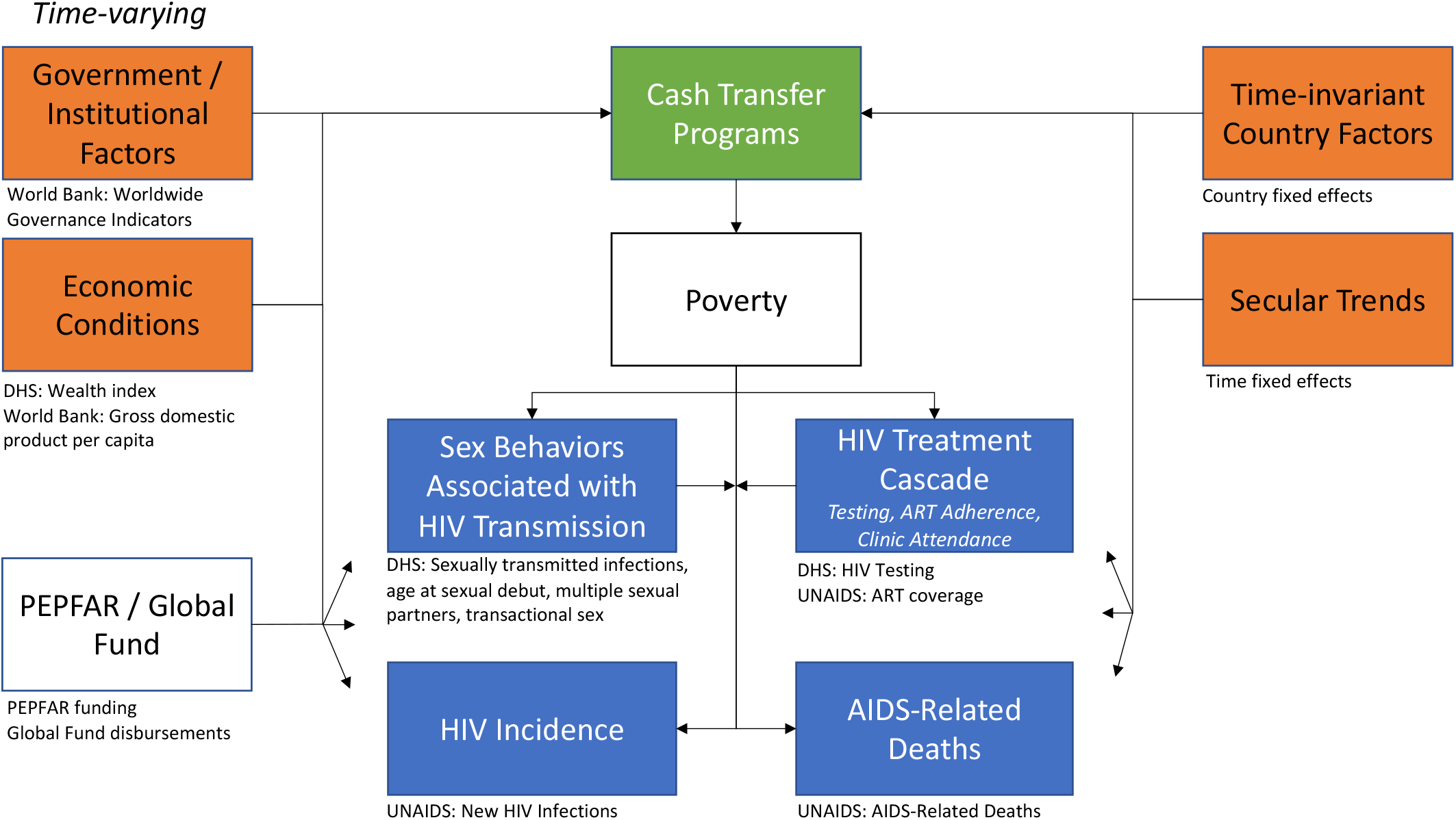
Proposed causal framework with directed acyclic graph (DAG) outlining potential relationships between large-scale cash transfer programs and HIV-related outcomes, mediated through an anti-poverty effect. The green box is the exposure of interest (cash transfer programs). The blue boxes are the HIV-related outcomes of interest, two more proximal (sex behaviors, HIV treatment cascade) and two more distal (HIV incidence, AIDS-related deaths). The orange boxes are ancestors of both the exposure and the outcomes (i.e., confounders). Underneath each box are the covariates used to measure the constructs within the boxes. Covariates from the DHS are individual-level, and all other covariates are country-level.

We examined the association between cash transfer programs and both individual- and country-level outcomes. For individual-level outcomes, the unit of observation was a surveyed person in a given country during a given year, and we stratified individual-level outcomes by sex.

Individual-level outcomes included the continuous variable age at sexual debut among youths and the binary variables sexually transmitted infection within the prior 12 months, greater than one sexual partner within the prior 12 months, HIV test within the prior 12 months, transactional sex within the prior 12 months, and condom use during the last sexual encounter. The transactional sex outcome was only analyzed for males because this question was only recently added to the female questionnaire in the DHS and there were not enough observations for meaningful comparisons.

For country-level outcomes, the unit of observation was the country-year. Country-level outcomes included the number of new HIV infections, the number of AIDS-related deaths, and the proportion of people with HIV receiving antiretroviral therapy.

We estimated linear regression models for continuous outcomes, logistic regression models for binary outcomes, and negative binomial regression models for outcomes aggregated as counts. We included fixed effects for each country, which adjusted for measured and unmeasured time-invariant differences between countries, and for each year, which controlled for secular trends in the outcomes across all countries. We used robust standard errors clustered at the country level. For all outcomes, we included additional time-varying, country-level covariates of GDP per capita, PEPFAR funding per capita, HIV-related disbursements by The Global Fund per capita, and three World Bank Worldwide Governance Indicators (Control of Corruption, Political Stability and Absence of Violence, and Voice and Accountability). The other three World Bank Worldwide Governance Indicators (Government Effectiveness, Regulatory Quality, and Rule of Law) were not included because of collinearity (Supplementary Appendix). For individual-level outcomes, we included additional covariates – age, single marital status, education, wealth quintile, and rural/urban household setting – and used survey commands to apply sampling probability weights.

We performed several secondary and sensitivity analyses to better characterize the association between cash transfer programs and HIV-related outcomes. First, because country-level outcomes were available annually, we evaluated the temporal relationship between cash transfer programs and country-level outcomes by creating a series of binary indicators for each year after the cash transfer period began. Second, we explored whether there were interactions between cash transfer programs and having above-median HIV prevalence (>3.7%) at the start of the cash transfer program. Third, we did a similar interaction analysis based on whether a country’s cash transfer program had above-median coverage (>23% of the population living below the poverty line). Fourth, we stratified models for individual-level outcomes by wealth quintile. Fifth, while our models controlled for PEPFAR and Global Fund spending, to further ensure there was no major collinearity contributing to our findings we used PEPFAR funding per capita and HIV-related Global Fund disbursements per capita as outcomes in our primary models to assess for correlation with cash transfer programs. Sixth, we assessed whether individual countries might be outliers for key outcomes by assessing whether estimates changed substantially after excluding each individually.

The difference-in-differences design is quasi-experimental and relies on the parallel trends assumption, which is that in the absence of the implementation of cash transfer programs, trends in outcomes would be similar in the intervention and comparison countries. We tested whether intervention and comparison countries had similar trends in the pre-cash transfer period by estimating regression models using only data prior to the cash transfer period in each country and including an interaction term between an indicator of whether the country was in the intervention group and a linear time trend. We tested the parallel trends assumption for outcomes with significant findings in our primary analysis. We further evaluated pre-trends in the country-level outcomes by including binary indicators for the four years prior to the cash transfer period in the previously mentioned temporal analysis. Using a temporal analysis to visualize pre-trends for the individual-level outcomes (which were measured in survey) is more difficult because annual survey data were not available for countries. As a result, sample sizes vary greatly by year. We attempted to mitigate this issue somewhat by categorizing multiple years together to allow for greater interpretability, but temporal trends for the individual-level outcomes should be interpreted with caution.

Recent advances in difference-in-differences analyses with variation in intervention timing have shown that estimates may be biased particularly if there is heterogeneity in intervention effect over time.^41,42^ We conducted a series of additional analyses to assess for the presence and magnitude of this potential bias,^43^ detailed in the Supplementary Appendix.

Additional details on the regression models are available in the Supplementary Appendix. We performed statistical analysis using SAS V.9.4 and R V.3.5.2 using the ggplot2 package.

### Data Availability Statement

Analyzed data can be requested from the DHS program website (https://www.dhsprogram.com/Data/) or are publicly available from UNAIDS (http://aidsinfo.unaids.org/), The World Bank (https://data.worldbank.org/data-catalog/), PEPFAR (https://data.pepfar.gov/financial), and The Global Fund (https://data-service.theglobalfund.org/downloads).

### Code Availability Statement

Analysis code is available upon request to the corresponding author.

## Results

Forty-two countries were eligible for inclusion in this study — 36 (86%) in Africa, 4 (10%) in Latin America and the Caribbean, and 2 (5%) in Asia (Table 1, Supplementary Figures 1-3). Among these, 21 countries implemented an eligible cash transfer program (or combination of cash transfer programs) at some point during the study period (Figure 1). In these countries, there were 36 cash transfer programs — 28 were unconditional and eight were conditional (Supplementary Table 1). The median total coverage level for cash transfer programs in the intervention group was 23% of the impoverished population (IQR 14-63%) and the median HIV prevalence at the beginning of the cash transfer period was 3.7% (IQR 1.5-10.7%).

**Table 1.** Characteristics of included countries that implemented a cash transfer program (or combination of programs) with greater than 5% impoverished population coverage during the study period (1996-2019) compared to those that did not implement such program(s).

|  | **Intervention Countries**<br>**N=21** | **Comparison Countries**<br>**N=21** | **Total**<br>**N=42** | **p-value** |
| --- | --- | --- | --- | --- |
| **Population (1000s), 1996, median (IQR)** | 10,372 (2,786-21,032) | 4,349 (1,663-7,251) | 11,801 (2,948-25,876) | 0.45 |
| **Region, N(%)** |  |  |  | <.0001 |
| *Africa* | 16 (76) | 20 (95) | 36 (86) |  |
| *Latin America / Caribbean* | 3 (14) | 1 (5) | 4 (10) |  |
| *Asia* | 2 (10) | 0 (0) | 2 (5) |  |
| **HIV Prevalence, median (IQR)** |  |  |  |  |
| *1996* | 4.1 (2.0-12.5) | 1.7 (1.4-4) | 2.8 (1.6-6.3) | 0.007 |
| *2005* | 4.4 (1.7-12.0) | 2.2 (1.4-4.0) | 2.7 (1.4-6.2) | 0.02 |
| *2019* | 3.2 (1.1-12.1) | 2.0 (1.3-3.4) | 2.4 (1.2-6.1) | 0.04 |
| **Annual HIV Incidence per 100,000, median (IQR)** |  |  |  |  |
| *1996* | 379 (160-988) | 218 (128-377) | 246 (145-484) | 0.01 |
| *2005* | 237 (77-539) | 130 (90-209) | 153 (86-350) | 0.06 |
| *2019* | 80 (26-273) | 59 (40-106) | 66 (38-172) | 0.32 |
| **Annual AIDS-related Death Rate per 100,000, median (IQR)** |  |  |  |  |
| *1996* | 137 (69-284) | 87 (45-113) | 94 (46-193) | 0.01 |
| *2005* | 168 (83-546) | 125 (73-147) | 132 (74-272) | 0.02 |
| *2019* | 47 (24-95) | 38 (22-79) | 47 (22-82) | 0.27 |
| **Proportion of population receiving ART, median (IQR)** |  |  |  |  |
| *2005* | 4 (3-7) | 2 (1-6) | 3 (2-7) | 0.09 |
| *2019* | 74 (62-82) | 57 (43-64) | 64 (46-79) | 0.01 |
| **PEPFAR recipient, N (%)** | 16 (76) | 8 (38) | 24 (57) | 0.03 |
| **PEPFAR funding per capita, median (IQR)** |  |  |  |  |
| *2005* | 0.0 (0.0-2.8) | 0.0 (0.0-0.7) | 0.0 (0.0-2.4) | 0.57 |
| *2019* | 6.9 (0.3-9.2) | 0.0 (0.0-3.2) | 1.0 (0.0-8.2) | 0.05 |
| **HIV Global Fund recipient, N (%)** | 21 (100) | 21 (100) | 42 (100) | n/a |
| **HIV Global Fund disbursements per capita, median (IQR)** |  |  |  |  |
| *2005* | 0.7 (0.2-1.6) | 0.7 (0.3-1.1) | 0.7 (0.2-1.6) | 0.4 |
| *2019* | 1.4 (1.1-3.1) | 1.4 (0.8-2.0) | 1.4 (0.8-2.7) | 0.13 |
| *World Bank Governance Indicators, 1996* |  |  |  |  |
| **Corruption, median (IQR)** | -0.7 (-1.1-0.1) | -0.6 (-0.9 - -0.1) | -0.7 (-1.0 – 0.0) | 0.68 |
| **Stability and Violence, median (IQR)** | -0.4 (-0.9 - -0.1) | -0.3 (-1.2 – 0.3) | -0.4 (-1.1 – 0.1) | 0.75 |
| **Voice and Accountability, median (IQR)** | -0.6 (-0.9 – 0.3) | -0.9 (-1.3 - -0.2) | -0.7 (-1.1 - -0.1) | 0.19 |
| **Effectiveness, median (IQR)** | -0.7 (-1.0 - -0.2) | -0.7 (-1.2 - -0.2) | -0.7 (-1.1 - -0.2) | 0.34 |
| **Rule of Law, median (IQR)** | -0.7 (-1.0 - -0.2) | -0.8 (-1.3 – 0.0) | -0.8 (-1.3 - -0.2) | 0.56 |
| **Regulatory Quality, median (IQR)** | -0.3 (-1.0 – 0.1) | -0.7 (-1.3 - -0.3) | -0.5 (-1.1 - -0.2) | 0.21 |

At the start of the study period, intervention countries had higher HIV prevalence (median 4.1% vs 1.7%) and annual HIV incidence rate (median 3.8 vs 2.2 per 1,000 persons) relative to comparison countries, but there was no difference between them in any of the six World Bank Governance Indicators (Table 1). All countries received some HIV-related Global Fund disbursements during the study period, and 16 (76%) intervention countries relative to 8 (38%) control countries received PEPFAR funding at some point during the study period.

We obtained individual survey data from 99 DHS and 6 AIS conducted in included countries during the study period – 24 during intervention years and 82 during comparison years (Figure 1). There were 1,885,733 survey respondents in total, of whom 1,295,177 (69%) were female and 545,867 (29%) were interviewed during intervention years (Supplementary Tables 4-6).

In our primary individual-level analyses, among females, cash transfer programs were associated with a lower probability of having a sexually transmitted infection within the last 12 months (odds ratio [OR] 0.67, 95% confidence interval [CI] 0.50-0.91) and higher probability of having had an HIV test within the last 12 months (OR 2.61, 95% CI 1.15-5.88) (Table 2, Supplementary Tables 7-17). PEPFAR funding per capita (OR 1.14 per $5 increase, 95% CI 1.01-1.30) and HIV-related Global Fund disbursements per capita (OR 1.48 per $5 increase, 95% CI 1.18-1.84) were also associated with an increased probability of having had an HIV test within 12 months.

**Table 2.** The relationship between cash transfer programs and individual- and country-level HIV-related outcomes.

| Outcomes | Cash Transfer Program | | PEPFAR Funding per capita (per \$5 increase) | | HIV-related Global Fund Disbursements per capita (per \$5 increase) | |
| --- | --- | --- | --- | --- | --- | --- |
|  | Effect Measure | 95% CI | Effect Measure | 95% CI | Effect Measure | 95% CI |
| *Individual-level, Females* |  |  |  |  |  |  |
| Age at sexual debut among youths, coefficient<sup>1</sup> | 0.00 | -0.09-0.10 | 0.03 | -0.01-0.07 | 0.01 | -0.09-0.12 |
| Sexually transmitted infection within 12 months, odds ratio<sup>1</sup> | 0.67 | 0.50 – 0.91 | 0.98 | 0.80-1.19 | 0.90 | 0.74-1.09 |
| Greater than 1 sexual partner within 12 months, odds ratio<sup>1</sup> | 1.04 | 0.75 – 1.46 | 1.17 | 0.89-1.54 | 0.88 | 0.67-1.15 |
| HIV test within 12 months, odds ratio<sup>1</sup> | 2.61 | 1.15 – 5.88 | 1.14 | 1.01-1.30 | 1.48 | 1.18-1.84 |
| Condom use at last sex, odds ratio<sup>1</sup> | 0.94 | 0.77-1.14 | 1.01 | 0.91-1.09 | 1.16 | 0.99-1.37 |
| *Individual-level, Males* |  |  |  |  |  |  |
| Age at sexual debut among youths, coefficient<sup>1</sup> | -0.14 | -0.28-0.01 | 0.04 | -0.04-0.11 | -0.023 | -0.16-0.12 |
| Sexually transmitted infection within 12 months, odds ratio<sup>1</sup> | 1.10 | 0.85 – 1.43 | 1.01 | 0.90-1.13 | 1.02 | 0.80-1.31 |
| Greater than 1 sexual partner within 12 months, odds ratio<sup>1</sup> | 1.12 | 0.99 – 1.28 | 1.02 | 0.91-1.13 | 1.00 | 0.91-1.09 |
| HIV test within 12 months, odds ratio<sup>1</sup> | 3.19 | 2.45 – 4.15 | 1.22 | 1.12-1.32 | 1.22 | 1.09-1.36 |
| Condom use at last sex, odds ratio<sup>1</sup> | 0.88 | 0.75 – 1.04 | 1.02 | 0.96-1.07 | 1.00 | 0.89-1.14 |
| Transactional sex within 12 months, odds ratio<sup>1</sup> | 0.99 | 0.85 – 1.15 | 0.93 | 0.80-1.09 | 1.07 | 0.88-1.31 |
| *Country-level* |  |  |  |  |  |  |
| New HIV infections, incidence rate ratio<sup>2</sup> | 0.94 | 0.89 – 0.99 | 1.00 | 0.99-1.01 | 0.99 | 0.98-1.00 |
| AIDS-related deaths, incidence rate ratio<sup>2</sup> | 0.99 | 0.95 – 1.03 | 0.98 | 0.97-0.99 | 0.99 | 0.98-1.01 |
| Proportion of people with HIV receiving ART, coefficient<sup>2</sup> | 5.0 | -0.2 – 10.1 | 2.6 | 1.7-3.5 | 3.3 | 0.4-6.2 |

<sup>1</sup> Multivariable models include cash transfer program, age, single marital status, education, wealth quintile, rural household setting, and the country-level covariates GDP per capita, PEPFAR funding per capita, HIV-related Global Fund disbursements per capita, and three World Bank Governance Indicators: Corruption, Stability and Violence, and Voice and Accountability.
<sup>2</sup> Multivariable models include the country-level covariates GDP per capita, PEPFAR funding per capita, HIV-related Global Fund disbursements per capita, and three World Bank Governance Indicators: Corruption, Stability and Violence, and Voice and Accountability.

Among males, cash transfer programs were significantly associated with an increased probability of having an HIV test within the last 12 months (OR 3.19, 95% CI 2.45-4.15) (Table 2, Supplementary Tables 7-17). PEPFAR funding per capita (OR 1.22 per $5 increase, 95% CI 1.12-1.32) and HIV-related Global Fund disbursements per capita (OR 1.22 per $5 increase, 95% CI 1.09-1.36) were also associated with an increased probability of having an HIV test within the last 12 months.

In our primary country-level analyses, cash transfer programs were associated with a reduction in new HIV infections (incidence rate ratio [IRR] 0.94, 95% CI 0.89-0.99), but not with the proportion of people with HIV receiving antiretroviral therapy (5.0%, 95% CI -0.2-10.1) or AIDS-related deaths (IRR 0.99, 95% CI 0.95-1.03). In the same models, PEPFAR funding per capita was associated with a reduction in AIDS-related deaths (IRR 0.98 per $5 increase, 95% CI 0.97-0.99) and an increase in the proportion of people with HIV receiving antiretroviral therapy (2.6% per $5 increase, 95% CI 1.7-3.5), results that are consistent with an earlier analysis of the relationship between PEPFAR and HIV outcomes.^44^ PEPFAR funding per capita was not associated with new HIV infections (IRR 1.00 per $5 increase, 95% CI 0.99-1.01). In addition, HIV-related Global Fund disbursements per capita were associated with an increase in the proportion of people with HIV receiving antiretroviral therapy (3.3% per $5 increase, 95% CI 0.4-6.2), but not with new HIV infections (IRR 0.99 per $5 increase, 95% CI 0.98-1.00) or AIDS-related deaths (IRR 0.99 per $5 increase, 95% CI 0.98-1.01).

We next evaluated associations between cash transfer programs and country-level outcomes over time (Figure 2). In fully adjusted models, we found that new HIV infections were significantly lower in the first year of the cash transfer program (IRR 0.94, 95% CI 0.89-0.99). While the effects on new infections became larger in subsequent years after the introduction of cash transfer programs, they were less precisely estimated over time as a result of declining numbers of observations and were no longer significant after the second year of the cash transfer program. There were no significant changes in AIDS-related deaths during the first year of the cash transfer program (IRR 0.99, 95% CI 0.95-1.03), consistent with our primary analysis. However, we found significant reductions in AIDS-related deaths by the second year of the cash transfer program (IRR 0.91, 95% CI 0.83-0.99), with larger reductions over time that peaked in the ninth year of the cash transfer program (IRR 0.75, 95% CI 0.57-0.99). Similarly, there was no significant change in the proportion of people with HIV receiving antiretroviral therapy on the first year of the cash transfer program (0.8%, 95% CI -1.0%-2.5%), but we found a significant increase by the second year (3.0%, 95% CI 0.3-5.7), with larger increases over time.

**Figure 2.**
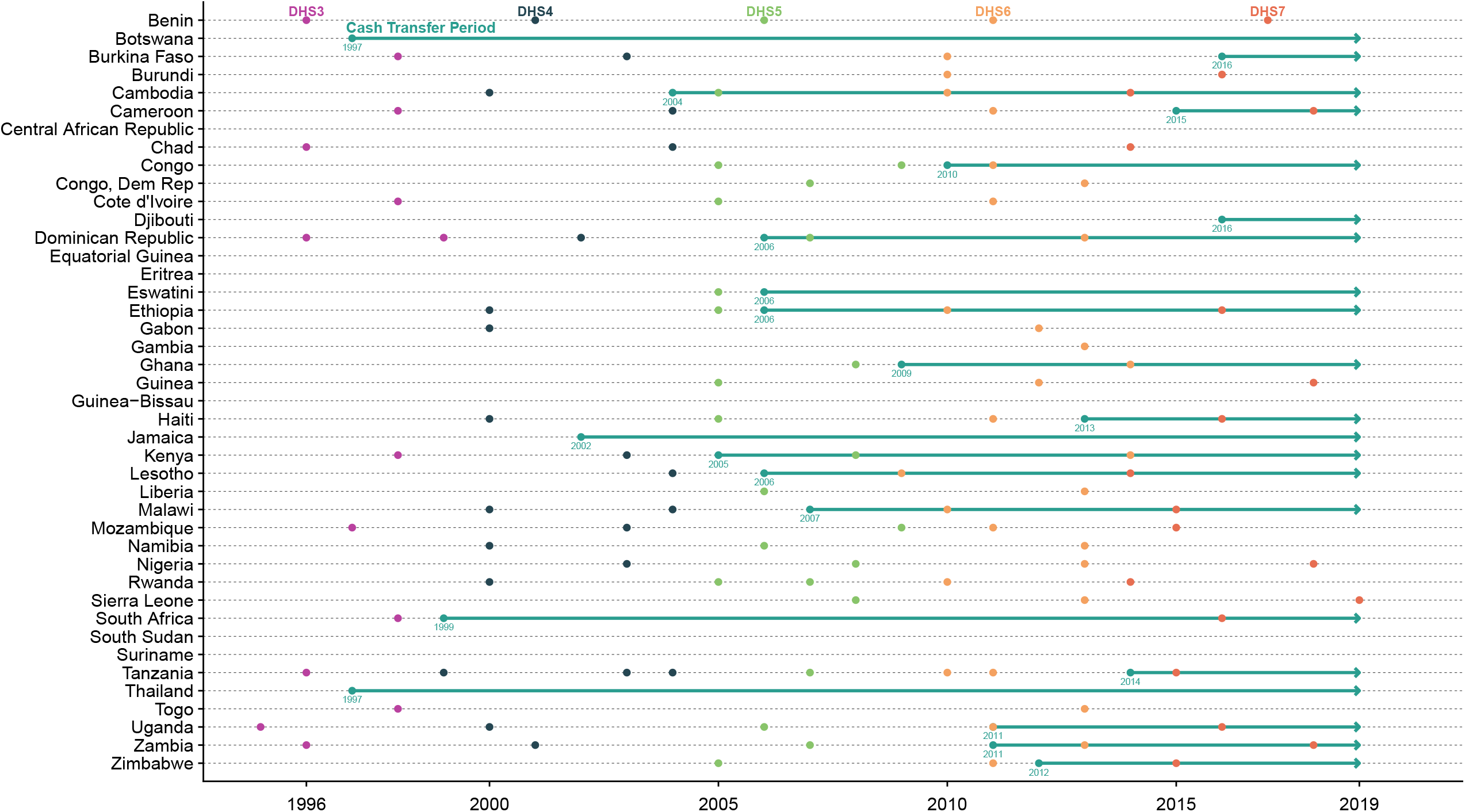
Timeline of included countries, with the green line indicating the cash transfer period, and the colored dots indicating years during which a Demographic and Health Survey (DHS) was conducted.

In the interaction analysis, the effects of cash transfer programs were greater in higher prevalence countries for the outcomes of an HIV test in the last 12 months among females (p<0.0001) and males (p<.0001), and in lower prevalence countries for new HIV infections (p=0.007) (Figure 3, Supplementary Table 21). The effects of cash transfer programs were greater with higher coverage cash transfer programs for the outcomes of sexually transmitted infection in the last 12 months among females (p<0.0001), having had an HIV test in the last 12 months among females (p=0.05) and males (p<.0001), and AIDS-related deaths (p=0.01).

**Figure 3.**
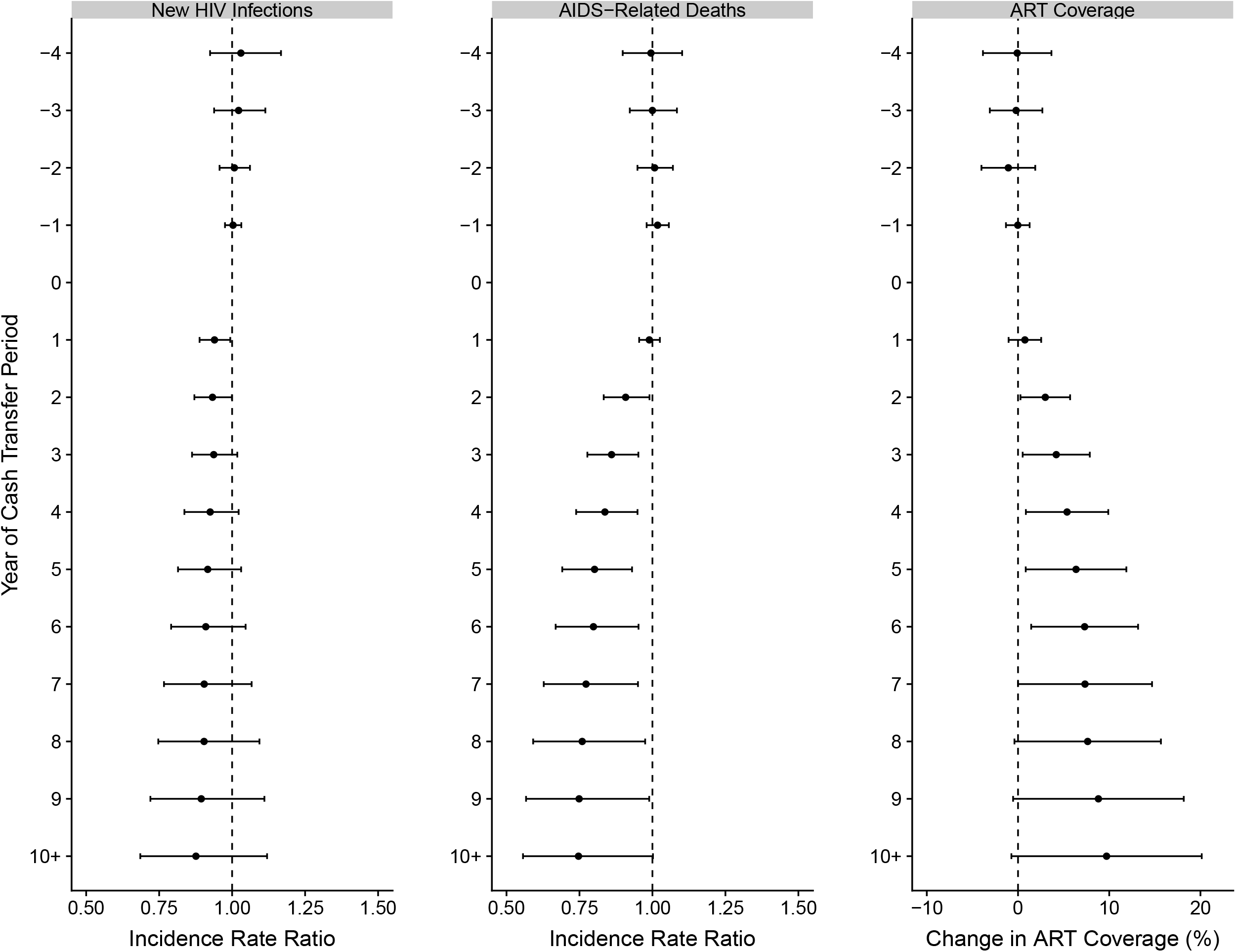
Adjusted incidence rate ratios of new HIV infections and AIDS-related deaths, and adjusted change in the proportion of proportion of people with HIV receiving antiretroviral therapy, as a function of year of the cash transfer period.

**Figure 4.**
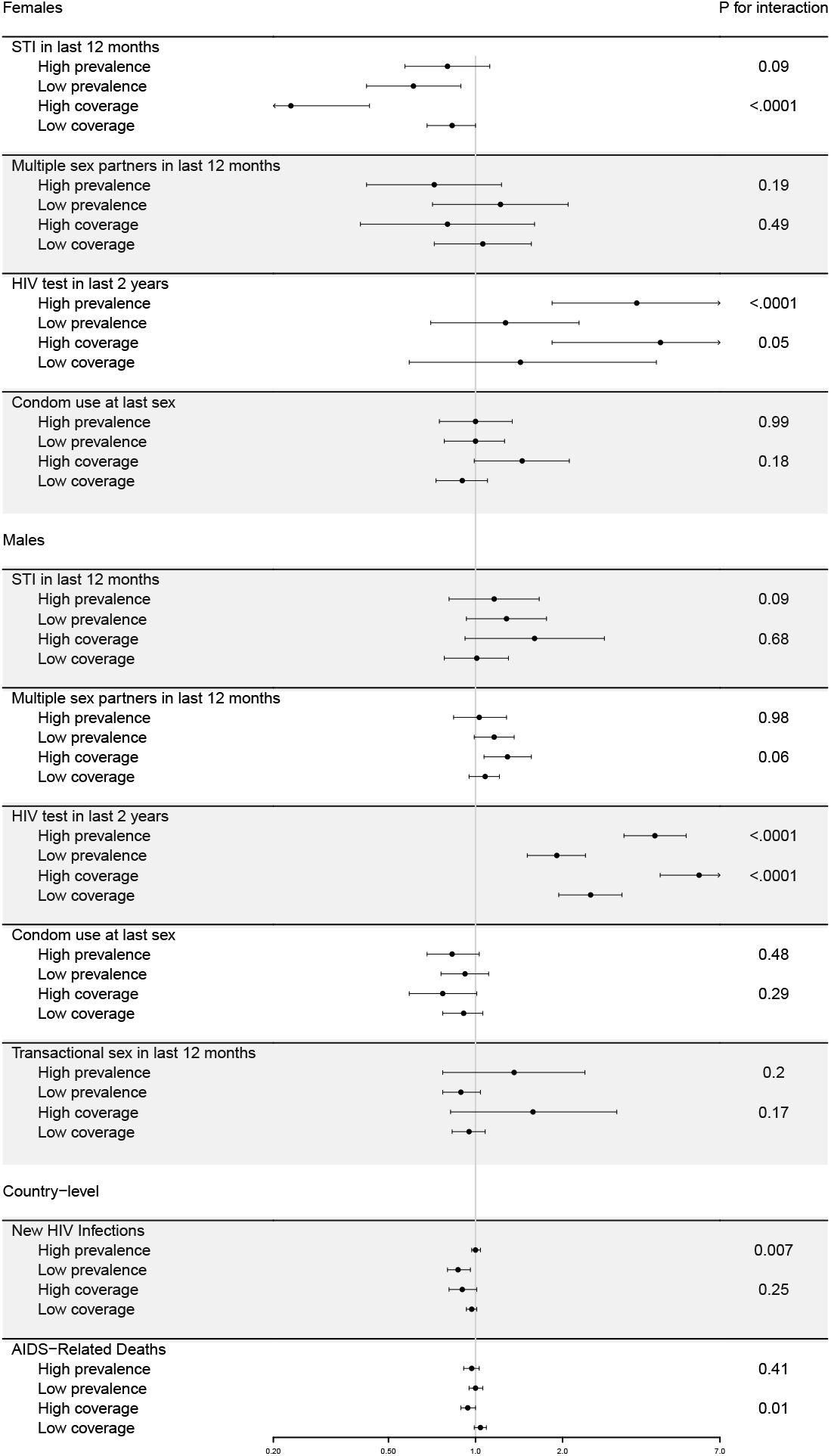
Interaction analysis of baseline HIV prevalence at the start of the cash transfer period (above or below the median, 3.7%) and impoverished population coverage of the cash transfer program(s) (above or below the median, 23%) for individual- (stratified by sex) and country-level outcomes, with adjusted odds ratios for individual-level outcomes and adjusted incidence rate ratios for country-level outcomes, and p-values for interaction.

When stratifying individual-level outcomes by wealth quintile, there were some modest trends suggesting larger effects in poorer segments of the population, though these were inconsistent and not definitively identified (Supplementary Figure 4). We confirmed that there was no significant association between the presence of cash transfer programs and either PEPFAR funding per capita or HIV-related Global Fund disbursements when these were included in our primary model as outcomes (Supplementary Tables 22-23). Exclusion of individual countries did not reveal possible outlier countries for any outcome except for HIV testing in females, for which Guinea and Zambia were potentially outliers whose exclusion substantially changed the estimated effect of cash transfers (Supplementary Tables 24-27).

Our fully adjusted models to test whether intervention and comparison countries had similar trends in outcomes before the introduction of cash transfers in a given country showed no differences between countries for the individual-level outcome of having a sexually transmitted infection in the last 12 months for females (OR 0.98, 95% CI 0.92-1.04) or males (OR 0.99, 95% CI 0.95-1.02), or for the country-level outcome of new HIV infections (IRR 0.99, 95% CI 0.96-1.02) (Supplementary Tables 28-30). There were small, significant differences of opposite magnitude in trends in outcomes before the introduction of cash transfers for the individual-level outcome of having an HIV test within the prior 12 months for females (OR 0.81, 95% CI 0.81-0.82) and males (OR 1.27, OR 1.01-1.12) (Supplementary Tables 31-32). There were no discernible differences in outcomes between intervention and control countries in the four years prior to the cash transfer period in our temporal analysis of country-level (Figure 2) or individual-level outcomes (Supplementary Figures 5-6) except for the HIV testing outcome among males, where there was some visual evidence of differential pre-trends in cash transfer countries.

Additional analyses suggested that the effect of cash transfers was heterogeneous over time but that any resultant bias was likely small (Supplementary Appendix, Supplementary Figures 7-8, Supplementary Tables 33-34).

## Discussion

In this study of 42 countries with generalized HIV epidemics of varying magnitude across three continents from 1996 to 2019, we found that sizeable cash transfer programs were associated with important improvements in HIV-related outcomes at both the population and individual levels. These included an immediate reduction in new HIV infections and delayed improvements in both AIDS-related deaths and the proportion of people with HIV receiving antiretroviral therapy, with benefits that grew larger over time. Among individuals, we found that cash transfer programs were associated with a reduction in sexually transmitted infections (a key proxy measure for risk of HIV transmission) among females, as well as large increases in recent HIV testing among males and females, though there were small differential pre-trends for the HIV testing outcome so this finding should be interpreted with some caution. Our interaction analyses showed that cash transfer programs with greater numbers of beneficiaries had the largest effects on HIV-related outcomes, suggesting an element of dose-response at the population level. We also found that the relationship between cash transfer programs and HIV testing was strongest in countries with higher baseline HIV prevalence, indicating the importance of the specific context of a given country’s HIV epidemic. To our knowledge, this is the first study to have combined data from all countries with generalized HIV epidemics and studied the effects of large-scale cash transfer programs.

While our findings are consistent with prior evidence from randomized controlled trials of cash transfer interventions that support the use of cash transfers for the prevention of HIV,^20,45-47^ and along the HIV care continuum,^25-34^ there are several notable distinctions to consider when interpreting our findings. First, the cash transfer programs considered in this study were generally of larger scale and less HIV-specific than those studied in the randomized trials.

Second, almost all of the cash transfer interventions studied in randomized trials were conditional on intermediary outcomes like school attendance, negative testing for sexually transmitted infections, HIV testing, or clinical follow up, whereas nearly 80% of the programs considered in our analysis were unconditional. Our study thus provides evidence, across many countries with generalized HIV epidemics, on the effects of primarily government-led cash transfer programs. Third, by evaluating outcomes for entire populations (i.e. by including individuals and households that did not receive transfers), our findings also capture the indirect or spillover effects of these interventions. These spillover effects are likely to be important in the context of an infectious disease with transmission dynamics and clinical outcomes that are heavily influenced by structural factors like poverty and food insecurity.

There are a number of hypothesized mechanisms by which cash transfer interventions could improve HIV-related outcomes. By increasing economic well-being, empowerment among women, and educational attainment, cash transfers may lead to lower risk sexual behaviors (as evidenced in our analysis by a reduction in sexually transmitted infections), thus lowering the probability of acquiring or transmitting HIV.^13^ This plausibly includes a reduction in transactional sex among women,^48^ an important driver of HIV risk among adolescent girls and young women in particular for which data were unavailable to consider in our analysis.^5^ Cash transfer programs may also lead to improvements along the HIV care continuum (i.e., HIV testing, clinic attendance, and antiretroviral adherence) through a direct economic mechanism that reduces barriers to care and a psychological mechanism that promotes health-seeking behaviors through improvements in mental bandwidth.^14^ As a result, cash transfers may lead to increases in HIV diagnoses (as evidenced in our analysis by increased HIV testing), engagement in clinical care by people with HIV, and higher probability of receiving and adhering to antiretroviral therapy with subsequent virologic suppression (as evidenced in our analysis by a delayed increase in the proportion of people with HIV receiving antiretroviral therapy). This would both directly improve clinical outcomes for people with HIV and reduce rates of transmission because of the highly effective strategy of using HIV treatment as prevention, commonly referred to as “Undetectable = Untransmittable” or “U=U.” By supporting preventive health behaviors, anti-poverty interventions like cash transfers can thus play an important role in improving individual HIV outcomes and preventing HIV transmission by intervening proximally to current efforts for HIV control, which are focused primarily at the health system level.

While previous analyses have used a similar design to study effects of programs like PEPFAR, this study is the first to do so for a common anti-poverty program that a growing number of LMICs are introducing as central features of their poverty reduction and social protection strategies.^44^ While not the primary objective of this study, our findings also suggest improvements in HIV testing, population antiretroviral coverage, and AIDS-related deaths related to PEPFAR and The Global Fund.

This study has several limitations. The cash transfer programs we considered were heterogeneous in terms of target population, size of transfer, conditionality, and coverage. Due to sample size limitations, we cannot precisely determine the relative importance of these other features of cash transfer programs, although in our interaction analyses we do establish that programs which covered more individuals tended to have larger effects. In particular, the relative amount of the transfer is likely to influence any effect it has on health outcomes, but because of variability of transfer size within many of the programs and inconsistent reporting we were unable to meaningfully consider this in our analysis. While the DHS and AIS surveys do not uniformly indicate whether participants received cash transfers, and thus we cannot separately determine cash transfer program effects on beneficiaries and non-beneficiaries, our objective was to evaluate the overall population-level effect of these programs, and it is plausible that the effects are larger on beneficiaries relative to non-beneficiaries. While we included country and year fixed effects and used a difference-in-differences design, the possibility of residual confounding related to unmeasured time-varying variables remains, though the robustness of our results after controlling for the available time-varying country-specific variables suggests this bias, if present, is minor. Specifically, there are country-specific policies that influence cash transfer program coverage and uptake (e.g., outreach, enrollment procedures, ease of benefit receipt). We attempted to control for these differences by including country fixed effects and the World Bank Governance Indicators in regression models, but if these policies differed between countries over time and were also associated with changes in HIV-related outcomes this may influence our findings. Importantly, though, we were attempting to examine the effects of cash transfer programs as they are delivered in the real world and would emphasize that implementation failures would most likely bias our results towards the null. The study period we considered was one of generally substantial expansion of HIV control programs, and the relationship between cash transfer programs and HIV-related outcomes may differ in settings with already established HIV care systems.

## Conclusion

In this difference-in-differences study of 42 countries with generalized HIV epidemics from 1996 to 2019, we found that cash transfer programs were associated with an immediate reduction in new HIV infections, a delayed improvement in AIDS-related deaths and the proportion of people with HIV receiving antiretroviral therapy, a reduction in sexually transmitted infections in the last 12 months among females, and an increase in recent HIV testing among males and females. Based on our results, experimental studies that further investigate the effects of unconditional cash transfers on HIV incidence and other HIV prevention behaviors should be a priority for future research. This study also contributes to our understanding of the social determinants of health, and suggests that HIV-related benefits should be included in cost-benefit analyses of cash transfer programs. As countries expand cash transfer programs, particularly in the context of the COVID-19 pandemic, these findings suggest that anti-poverty interventions like cash transfers should receive greater attention as part of HIV control efforts, alongside the already existing focus on expanding biomedical services.

## Author Contribution Statement

AR and HT conceptualized and designed the study. AR conducted the primary analysis and wrote the first draft of the manuscript, both with critical feedback from HT.

## Abbreviations

CCT: Conditional cash transfer
CI: Confidence Interval
DHS: Demographic and Health Survey
IRR: incidence rate ratio
LMIC: low- and middle-income countries
OR: odds ratio
PEPFAR: President’s Emergency Plan for AIDS Relief
UCT: unconditional cash transfer
UNAIDS: The Joint United Nations Programme on HIV/AIDS

