## Supplemental Appendix for "Cash Transfer Programs and HIV-Related Outcomes: an Analysis of 42 Countries from 1996 to 2019"

**Supplementary Appendix**

### Identification of Cash Transfer Programs, Calculation of Impoverished Population Coverage, Definition of Primary Explanatory Variable

We identified all major cash transfer programs within included countries. Cash transfer programs were defined as non-contributory monetary transfers to individuals or households and included unconditional cash transfers, conditional cash transfers, social pensions and enterprise grants. We manually searched a variety of sources to identify the programs as well as the year in which they were implemented, target population, conditionality, amount of transfer, and the most recently available number of beneficiaries.^1-5^

For each cash transfer program, we calculated impoverished population coverage — program coverage as a proportion of the population with income less than the international poverty line ($1.90 per day in 2011 Purchasing Power Parity [PPP]). To account for spillover effects, if only an estimate for direct beneficiaries was available, we first multiplied direct beneficiaries by the average household size to estimate total beneficiaries.^6^ We then divided the most recent estimate of total beneficiaries (direct and indirect) by the impoverished population. We calculated the impoverished population by multiplying the poverty headcount (the percentage of the population with income less than the international poverty line) prior to program implementation by the mid-year population from the year of the total beneficiaries estimate.^7^ We used the poverty headcount prior to program implementation because cash transfer programs typically reduce poverty. As a result, using poverty headcount estimates after program implementation (which may have been decreased by the cash transfer programs) to calculate impoverished population coverage will not as accurately capture the impoverished population covered by the programs.

Our primary explanatory variable of interest was a binary variable indicating presence of a cash transfer program (or combination of programs) implemented during the study period with impoverished population coverage greater than 5%, and we defined the cash transfer period as the years during which this was the case. We chose 5% empirically as the smallest likely coverage with which we might expect to see population effects, with a pre-specified plan for secondary interaction analyses of greater degrees of impoverished population coverage. We considered the cash transfer program status as of the start of each year in our analysis (i.e. if the cash transfer period began in a given country in July, 2004, then 2004 would be coded as not being in the cash transfer period and 2005 would be coded as being in the cash transfer period).

There were 21 included countries that introduced a cash program (or combination of cash programs) with impoverished population coverage greater than 5% during the study period. In these intervention countries there were 36 cash transfer programs introduced during the study period, of which 28 were unconditional (Supplementary Table 1). We also identified 20 cash transfer programs introduced during the study period in control countries that did not reach a total of 5% impoverished population coverage (Supplementary Table 2), and 8 cash transfer programs introduced prior to the study period that were thus ineligible for inclusion in this analysis (Supplementary Table 3).

### Demographic and Health Survey Sampling, Ethics, and Wealth Index

Sampling methods for the Demographic and Health Surveys (DHS) have been previously described,^8^ but typically use a two-stage cluster sampling design to produce representative national and sub-national estimates for a variety of indicators. The first stage involves systematic selection of Enumeration Areas drawn from census files with probability proportional to population size, and the second stage involves a random sampling of households from each Enumeration Area.

Procedures and questionnaires for DHS surveys have been reviewed and approved by the Independent Consulting Firm Institutional Review Board, and all analyzed data were anonymized.

The wealth index is a composite measure of a household’s cumulative living standard calculated by the DHS using ownership of certain assets, materials used for housing construction, and types of water access and sanitation facilities. Through the wealth index, survey respondents are categorized into wealth quintiles.

### Statistical Analysis

#### Primary models for country-level outcomes

Country-level outcomes included the number of new HIV infections (available 1996-2019), the number of AIDS-related deaths (available 1996-2019), and the proportion of people with HIV receiving antiretroviral therapy (available 2000-2019). For these outcomes we estimated regression models with the following generic form:

**Y**_jt_ = α_j_ + β***C***_jt_ + δ***X***_jt_ + γ_t_ + ε_jt,_

where **Y**_jt_ was the outcome variable for country *j* during year *t*, α_j_ was a fixed effect for country *j* to control for time-invariant differences between countries, ***C***_jt_ was set to 1 if a combination of cash programs with impoverished population coverage >5% implemented during the study period was active in country *j* at the beginning of year *t*, **X**_jt_ was a vector of covariates for country *j* during year *t*, and γ_t_ was a fixed effect for year *t* to control for secular trends in the outcome.

The following covariates were included in ***X***: Gross Domestic Product (GDP) per capita, PEPFAR funding per capita as a continuous variable, HIV-related disbursements by The Global Fund per capita as a continuous variable, and three World Bank Worldwide Governance Indicators as continuous variables - Control of Corruption, Political Stability and Absence of Violence, and Voice and Accountability. We modeled PEPFAR funding and HIV-related Global Fund disbursements in $5 increments because this allowed interpretability and consistency across all outcomes and was close to the median value in intervention countries in 2019. The other three World Bank Worldwide Governance Indicators were considered for inclusion but ultimately left out of the models because they displayed multicollinearity with the other covariates as evidenced by variance inflation factors >5. There were no missing data in the country-level analysis during the years the outcomes were available, and we performed a complete case analysis.

The parameter of interest was β, which denotes the association between the presence of a cash transfer program and our outcomes. We used negative binomial regression models for outcomes aggregated as counts (number of new HIV infections, the number of AIDS-related deaths) and in these models included an offset variable (the natural log of mid-year population size for a given country and year). We used a linear regression for the continuous outcome (proportion of people with HIV receiving antiretroviral therapy). For the negative binomial models, we reported the exponentiated coefficient β as the incident rate ratio (IRR). For the linear regression model, we reported the coefficient of interest β, which represents the average change in the proportion of people with HIV receiving antiretroviral therapy after cash transfer program implementation. For all models, we reported 95% confidence intervals using robust standard errors with clustering at the country level.

#### Primary models for individual-level outcomes

Individual-level outcomes included the continuous variable age at sexual debut among youths and the binary variables sexually transmitted infection within the prior 12 months, greater than 1 sexual partner within the prior 12 months, HIV test within the prior 12 months, transactional sex within the prior 12 months, and condom use during the last sexual encounter. Outcomes were stratified by sex, and the transactional sex outcome was only evaluated for males because this question was only recently added to the female questionnaire. For these outcomes we estimated regression models with the following generic form:

**Y**_ijt_ = α_j_ + β***C***_jt_ + δ***X***_jt_ + *μ****Z***_i_ + γ_t_ + ε_ijt,_

where **Y**_ijt_ was the outcome variable for individual *i* in country *j* during year *t*, α_j_ was a fixed effect for country *j* to control for time-invariant differences between countries, ***C***_jt_ was set to 1 if a combination of cash programs with impoverished population coverage >5% implemented during the study period was active in country *j* at the beginning of year *t*, **X**_jt_ was a vector of covariates for country *j* during year *t*, ***Z***_i_ was a vector of covariates for individual *i*, and γ_t_ was a fixed effect for year *t* to control for secular trends in the outcome.

The following covariates were included in ***X***: GDP per capita, PEPFAR funding per capita as a continuous variable, HIV-related disbursements by The Global Fund per capita as a continuous variable, and three World Bank Worldwide Governance Indicators as continuous variables - Control of Corruption, Political Stability and Absence of Violence, and Voice and Accountability. We modeled PEPFAR funding and HIV-related Global Fund disbursements in $5 increments because this allowed interpretability and consistency across all outcomes and was close to the median value in intervention countries in 2019. The other three World Bank Worldwide Governance Indicators were considered for inclusion but ultimately left out of the models because they displayed multicollinearity with the other covariates as evidenced by variance inflation factors >5.

The following covariates were included in ***Z***: age as a continuous variable, single marital status as a binary variable, education as a categorical variable (none, primary, secondary, or greater than secondary), wealth quintile as a categorical variable (richest, richer, middle, poorer, or poorest), and rural setting as a binary variable. We performed a complete case analysis.

The parameter of interest was β, which denotes the association between the presence of a cash transfer program and our outcomes. We used logistic regression models for binary outcomes (sexually transmitted infection within the prior 12 months, greater than 1 sexual partner within the prior 12 months, HIV test within the prior 12 months, transactional sex within the prior 12 months, and condom use during the last sexual encounter). We used a linear regression for the continuous outcome (age at sexual debut among youths). For the logistic regression models, we reported the exponentiated coefficient β as the odds ratio (OR). For the linear regression model, we reported the coefficient of interest β, which represents the average change in the age of sexual debut among youths after cash transfer program implementation. For all models, we reported 95% confidence intervals using robust standard errors with clustering at the country level and used survey commands to apply sampling probability weights.

#### Association between cash transfer programs and study outcomes over time

We evaluated the temporal relationship between cash transfer programs and country-level outcomes, which were available on an annual basis. To do this, we estimated regression models with the following generic form:

**Y**_jt_ = α_j_ + χ***Cyear***_jt_ + δ***X***_jt_ + γ_t_ + ε_jt,_

where **Y**_jt_ was the outcome variable for country *j* during year *t.* Instead of ***C****_jt_*, we included ***Cyear***_jt_, a vector of binary variables indicating year of cash transfer program implementation (year -4, year -3, …, year 0, year 1, …, years 10+). The coefficients of interest were a vector of χ coefficients indicating the effect measure for the outcome as a function of time relative to the beginning of the cash transfer period.

We conducted a similar temporal analysis of the relationship between cash transfer programs and individual-level outcomes to evaluate pre-trends. However, because data were not available for countries during every year, sample sizes vary greatly by year. As a result, it was difficult to interpret trends when including a vector of binary variables by year, so instead we categorized multiple years together (years -7+, years -4-6, years -3-1, years 0-1, years 2-4, years 5-7, years 8+).

#### Interaction between cash transfer programs and baseline HIV prevalence

By introducing an interaction term to our regression models, we explored whether there was an interaction between cash transfer programs and baseline HIV prevalence at the beginning of the cash transfer period. For this analysis, we dichotomized intervention countries based on whether they had baseline HIV prevelance greater than the median value (3.7%).

For the country-level outcomes we estimated regression models with the following generic form:

**Y**_jt_ = α_j_ + κ***P****_medj_*****C***_jt_ + β***C***_jt_ + δ***X***_jt_ + γ_t_ + ε_jt,_

where Y_jt_ was the outcome variable for country *j* during year *t,* ***P****_medj_* was set to 1 if the HIV prevalence in country *j* at the start of the cash transfer period was greater than 3.7%.

We calculated the p-value for the interaction term (***P****_medj_*****C***_jt_) for all models. For each model we reported one effect measure and 95% confidence interval for low prevalence countries and one for high prevalence countries.

For the individual-level outcomes we estimated regression models with the following generic form:

**Y**_ijt_ = α_j_ + κ***P****_medj_*****C***_jt_ + β***C***_jt_ + δ***X***_jt_ + *μ****Z***_i_ + γ_t_ + ε_ijt,_

where **Y**_ijt_ was the outcome variable for individual *i* in country *j* during year *t,* and ***P****_medj_* was set to 1 if the HIV prevalence in country *j* at the time of cash transfer program implementation was greater than 3.7%.

We calculated the p-value for the interaction term (***P****_medj_*****C***_jt_) for all models. For each model we reported an effect measure and 95% confidence interval for low prevalence countries and one for high prevalence countries.

#### Interaction between cash transfer programs and impoverished population coverage

By introducing an interaction terms to our regression models, we explored whether there was an interaction between cash transfer programs and cash transfer program impoverished population coverage. For this analysis, we dichotomized intervention countries based on whether they had impoverished population coverage greater than the median value (23%).

For the country-level outcomes we estimated regression models with the following generic form:

**Y**_jt_ = α_j_ + ν***I****_medj_*****C***_jt_ + β***C***_jt_ + δ***X***_jt_ + γ_t_ + ε_jt,_

where **Y**_jt_ was the outcome variable for country *j* during year *t,* and ***I****_med_*_j_ was set to 1 if the impoverished population coverage in country *j* was greater than 23%.

We calculated the p-value for the interaction term (***I****_medj_*****C***_jt_) for all models. For each model we reported an effect measure and 95% confidence interval for low prevalence countries and one for high prevalence countries.

For the individual-level outcomes we estimated regression models with the following generic form:

**Y**_ijt_ = α_j_ + ν***I****_med_*_j_****C***_jt_ + β***C***_jt_ + δ***X***_jt_ + *μ****Y***_i_ + γ_t_ + ε_ijt,_

where **Y**_ijt_ was the outcome variable for individual *i* in country *j* during year *t,* and ***I****_med_*_j_ was set to 1 if the impoverished population coverage in country *j* was greater than 23%.

We reported the p-value for the interaction term (***I****_medj_*****C***_jt_) for all models. For each model we reported an effect measure and 95% confidence interval for low prevalence countries and one for high prevalence countries.

*Test of the parallel trends assumption*

We tested the parallel trends assumption by estimating regression models using only data prior to the implementation of cash transfer programs in each country and including an interaction term between an indicator of whether the country was in the intervention group (i.e. ultimately implemented cash transfer programs with impoverished population coverage greater than 5%) and a linear time trend. We tested the parallel trends assumption for the outcomes sexually transmitted infection in the last 12 months and number of new HIV infections because these were the individual- and country-level outcomes with significant findings and the greatest number of observations.

To do so, for the country-level outcome number of new HIV infections we estimated a regression model with the following generic form:

**Y**_jt_ = α_j_ + β***C***_j_ + σ***Year***_t_ + τ***C***_j_****Year***_t_ + δ***X***_jt_ + γ_t_ + ε_jt,_

where Y_jt_ was the number of new HIV infections for country *j* during year *t,* ***C***_j_ was set to 1 if a combination of cash programs with impoverished population coverage >5% was eventually implemented during the study period in country *j*, and ***Year***_t_ was a linear time trend. The coefficient of interest was τ, which showed whether pre-intervention trends were different in intervention countries compared to control countries.

To test the parallel trends assumption for the individual-level outcomes sexually transmitted infection and HIV testing in the last 12 months, we estimated regression models with the following generic form:

**Y**_ijt_ = α_j_ + β***C***_j_ + σ***Year***_t_ + τ***C***_j_****Year***_t_ + δ***X***_jt_ + *μ****Z***_i_ + γ_t_ + ε_ijt,_

where **Y**_ijt_ was the outcome variable for individual *i* in country *j* during year *t*, ***C***_j_ was set to 1 if a combination of cash programs with impoverished population coverage >5% was eventually implemented during the study period in country *j*, and ***Year***_t_ was a linear time trend. The coefficient of interest was τ, which showed whether pre-intervention trends were different in intervention countries compared to control countries.

#### Additional analyses to assess for treatment heterogeneity and degree of resultant bias

Recent advances in difference-in-differences analyses with variation in intervention timing have shown that estimates may be biased particularly if there is heterogeneity in intervention effect over time.^9,10^ This bias may arise when treatment effects are not homogenous and when treated units receive negative weights, which are proportional to the residuals from the regression model. To address this possibility we conducted a series of additional diagnostics and analyses proposed by Jakiela.^11^ To assess for possible treatment heterogeneity over time we used the ART coverage outcome because the effect appeared to change over time. To do so, we first plotted the residuals scaled by the sum of the squared residuals in Supplementary Figure 7, and showed that some treated country-year observations receive negative weight. Next, we evaluated the distribution of negative weights across intervention countries and over time (Supplementary Figure 8). In general, most country-years with negative weight tended to be during later program years. Taken together, this does suggest that the treatment effect may be heterogeneous over time. To assess for the magnitude of potential bias from this, we repeated our primary analyses after excluding country-years after year 4 of the cash transfer program (Supplementary Table 33), and after excluding countries that had negative weights during the early program years (Ghana, Jamaica, Dominican Republic, and Congo). There were no substantial changes in our primary findings with these additional analyses.

### Supplementary Table 1. Characteristics of identified cash transfer programs implemented within intervention countries during the study period (1996-2019), with total impoverished population coverage greater than 5%.

| Country | Program Name | First Complete Year Implemented | Conditionality and Target Population | Annual Transfer Amount (USD) | Beneficiaries Estimate | Total Beneficiaries | Poverty headcount ^7^ | Impoverished Population Coverage | Transfer Amount as % GDP per Capita |
| --- | --- | --- | --- | --- | --- | --- | --- | --- | --- |
| Botswana | Old Age Pension | 1997 | Unconditional, elderly | $360 | 108,870 (2018)^4^ | 446,367 | 33.3% (1993) | 59% | 4% |
| Burkina Faso | Burkin-Naong-Sa ya^2^ | 2016 | Unconditional, poor women with children | $224 | 908,537 (2018) | 908,537 | 43.8% (2014) | 12% | 12% |
| Cambodia | MoEYS scholarships for school children^12^ | 2004 | Conditional, school children | $60 | 156,519 (2017) | 719,987 | 17.7%^1^ (2012) | 22% | 4% |
| Cambodia | NOURISH^13^ | 2015 | Conditional, poor families with pregnant mothers or children | Varied | 5,554 (2016) | 25,548 | 17.7% (2012) | 1% | Varied |
| Cameroon | Social Safety Nets^5,14^ | 2015 | Unconditional, poor households | $360 | 634,756 (2020) | 634,756 | 26% (2014) | 10% | 11% |
| Congo | Fonds de Soutien à l’Agriculture (FSA)^4,15^ | 2010 | Unconditional, agricultural enterprise | Varied | 40,000 (2015)^4^ | 172,000 | 55.1% (2005) | 7% | Varied |
| Congo | Lisungi Safety Nets Project^2^ | 2015 | Conditional, households with children; Unconditional, elderly | $144-652 | 37,574 (2015) | 161,579 | 38.2% (2011) | 9% | 4-20% |
| Djibouti | Programme National de Solidarité Famille^5^ | 2016 | Unconditional, poor households | $680 | 91,526 (2015)^4^ | 91,526 | 22.3% (2013) | 45% | 24% |
| Dominican Republic | Programa Solidaridad^3^ | 2006 | Conditional, variety of target populations | Varied | 864,542 (2018)^4^ | 864,542 | 2.6% (2012) | 100% | Varied |
| Eswatini | Old Age Grant^5,16^ | 2006 | Unconditional, elderly and poor | $80 | 63,500 (2014)^4^ | 298,450 | 43% (2009) | 63% | 1% |
| Ethiopia | Productive Safety Net Programme (PSNP)^2,17^ | 2006 | Unconditional (cash and/or food), food insecure households | Equivalent of 15kg cereal, 4kg pulses | 10,000,000 (2015) | 46,000,000 | 39% (2004) | 100% | N/A |
| Ethiopia | Tigray Social Cash Transfer Pilot Programme (SCTPP)^2^ | 2012 | Unconditional, poor households | $372-804 | 17,705 (2014)^4^ | 17,705 | 35.2% (2010) | <0.5% | 31-66% |
| Ghana | Livelihood Empowerment Against Poverty (LEAP)^5^ | 2009 | Unconditional, elderly and disabled; Conditional, households with orphans and vulnerable children | $66-108 | 939,022 (2016)^4^ | 939,022 | 24.1% (2005) | 14% | 2-4% |
| Haiti | Ti Manman Cheri^3^ | 2013 | Conditional, children | $56 | 86,234 (2014) | 423,429 | 24.5% (2012) | 16% | 3% |
| Jamaica | Programme of Advancement Through Health and Education (PATH)^3^ | 2002 | Conditional,  Children, elderly, disabled, pregnant/ breastfeeding women, unemployed | Varied | 265,285 (2018)^4^ | 822,383 | 2.6% (1999) | 100% | Varied |
| Kenya | Cash Transfers for Orphans and Vulnerable Children (CT-OVC)^5^ | 2005 | Conditional and unconditional, poor households with at least one orphan or vulnerable child | $252 | 1,265,000 (2018)^4^ | 1,265,000 | 43.9% (2005) | 6% | 13% |
| Kenya | Older Persons’ Cash Transfer (OPCT)^5^ | 2007 | Unconditional, elderly | $264 | 310,000 (2017)^4^ | 1,116,000 | 43.9% (2005) | 5% | 12% |
| Kenya | Hunger Safety Net Programme (HSNP)^5^ | 2009 | Unconditional, food insecure households | $126 | 507,190 (2016)^4^ | 507,190 | 43.9% (2005) | 2% | 6% |
| Kenya | Persons with Severe Disability Cash Transfer (PWSD-CT)^5^ | 2011 | Unconditional, poor disabled | $216 | 27,200 (2013) | 106,080 | 43.9% (2005) | 0.5% | 9% |
| Lesotho | Old Age Pension^5,18^ | 2006 | Unconditional, elderly | $480 | 83,751 (2017)^4^ | 276,378 | 61.9% (2002) | 21% | 27% |
| Lesotho | Child Grants Programme (CGP)^5,18^ | 2010 | Unconditional, poor households with orphans or vulnerable children | $600 | 117,600 (2015)^4^ | 117,600 | 61.9% (2002) | 9% | 28% |
| Malawi | Mchinji Social Cash Transfer^5^ | 2007 | Unconditional, poor households | $66 | 782,561 (2016)^4^ | 782,561 | 73.9% (2004) | 6% | 8% |
| South Africa | Child Support Grant^5^ | 1999 | Unconditional, poor children | $270 | 11,703,165 (2015)^4^ | 47,982,976 | 36.3% (1996) | 100% | 4% |
| South Africa | Older Persons’ Grant^5^ | 2005 | Unconditional, elderly poor | $1200 | 3,086,851 (2015)^4^ | 10,186,608 | 34.8% (2000) | 53% | 12% |
| South Africa | Disability Grant^5^ | 2005 | Unconditional, disabled | $1200 | 1,098,018 (2015) | 3,623,459 | 34.8% (2000) | 19% | 12% |
| South Africa | Foster Child Grant^5^ | 2005 | Unconditional, foster children | $696 | 539,791 (2015) | 1,781,310 | 34.8% (2000) | 9% | 7% |
| South Africa | Grant In Aid^5^ | 2005 | Unconditional, for recipients who require regular attendance by another person | $264 | 119,541 (2015) | 394,485 | 34.8% | 2% | 3% |
| Tanzania | Community-Based Conditional Cash Transfer^5^ | 2011 | Conditional, children and vulnerable elderly | $72-432 | 13,000 (2013) | 13,000 | 60.3% | <0.5% | 3-19% |
| Tanzania | Productive Social Safety Net Programme^5^ | 2014 (massively scaled up) | Unconditional, poor households; Conditional, children / pregnant women | $276 | 5,164,623 (2016)^4^ | 5,164,623 | 49.6% | 20% | 12% |
| Thailand | Allowances for People Living with Disabilities^5^ | 1997 | Unconditional, disabled | $276 | 1,491,947 (2017) | 4,625,036 | 0% (2014) | N/A | 4% |
| Thailand | Allowances for People Living with HIV/AIDS^5^ | 2001 | Unconditional, people with HIV | $168 | 84,829 (2017) | 262,970 | 2.4% (2000) | 16% | 2% |
| Thailand | Child Support Grant^5^ | 2016 | Unconditional, poor families with children | $240 | 310,041 (2017) | 961,127 | 0% (2014) | N/A | 1% |
| Uganda | Social Assistance Grant for Empowerment (SAGE)^5^ | 2011 | Unconditional, elderly and vulnerable families | $96 | 150,000 (2018)^4^ | 675,000 | 44.5% (2009) | 4% | 4% |
| Uganda | Northern Uganda Social Action Fund II (Household Income Support Programme)^19,20^ | 2011 | Unconditional, demand-driven livelihood investments | Varied | 510,138 (2016)^4^ | 510,138 | 44.5% (2009) | 3% | Varied |
| Zambia | Social Cash Transfer Programme^5^ | 2011 | Unconditional, variety of target populations | $36 | 2,600,000 (2016)^4^ | 2,600,000 | 65.8% (2010) | 24% | 1% |
| Zimbabwe | Harmonised Social Cash Transfer (HSCT) | 2012 | Unconditional, poor households | $120-300 | 218,400 (2015)^4^ | 218,400 | 21.4% (2011) | 7% | 5-13% |

^1^ Cambodian national poverty line $0.98 2011 USD

### Supplementary Table 2. Characteristics of identified cash transfer programs implemented within control countries during the study period (1996 to 2019), with total impoverished population coverage less than or equal to 5%.

| Country | Program Name | First Complete Year Implemented | Beneficiaries Estimate | Total Beneficiaries | Poverty headcount^7^ | Impoverished Population Coverage |
| --- | --- | --- | --- | --- | --- | --- |
| Benin | Projet de Services Décentralisés Conduits par les Communautés (PSDCC)^2^ | 2016 | 13,000 (2015)^4^ | 71,500 | 49.6% (2015) | 1% |
| Burkina Faso | Nahouri^2^ | 2009 | 2,600 (2011) | 14,820 | 57.4% (2003) | <0.5% |
| Burundi | Terintambwe^4^ | 2013 | 10,000 (2013) | 10,000 | 78.6% (2006) | <0.5% |
| Burundi | Merankabandi^21^ | 2018 | 56,090 (2020) | 269,232 | 72.8% (2013) | 3% |
| Chad | Social Safety Nets^22^ | 2018 | 7,604 (2019) | 44,103 | 38% (2011) | 1% |
| Congo, Dem. Rep | UNICEF ARCC II^23^ | 2015 | 64,343 (2015)^4^ | 64,343 | 77.2% (2012) | <0.5% |
| Côte d’Ivoire | Programme National des Filets Sociaux Productifs^24^ | 2016 | 50,000 (2019)^4^ | 250,000 | 29.8% (2015) | 3% |
| Guinea | Productive Social Safety Net Programme^25^ | 2013 | 1,800 (2014) | 11,340 | 36.1% (2012) | <0.5% |
| Guinea | Cash Transfer for Health, Nutrition and Education^5^ | 2014 | 10,000 (2012)^4^ | 63,000 | 36.1% (2012) | 2% |
| Mali | Jigisemejiri (Tree of Hope)^5^ | 2014 | 321,790 (2016)^4^ | 321790 | 50% (2009) | 4% |
| Nigeria | In Care of the Poor (COPE)^5^ | 2008 | 22,000 (2007) | 101,200 | 56.3% (2003) | <0.5% |
| Nigeria | Ekiti State Social Security Scheme^5^ | 2012 | 25,000 (2013)^4^ | 115,000 | 56.4% (2009) | <0.5% |
| Nigeria | Subsidy Reinvestment and Empowerment Programme (SURE-P): Maternal and Child Health (MCH)^5^ | 2013 | 26,461 (2014) | 121,720 | 56.4% (2009) | <0.5% |
| Nigeria | Osun Elderly Persons Scheme^5^ | 2013 | 1,692 (2015) | 8,290 | 56.4% (2009) | <0.5% |
| Rwanda | Rwanda Demobilization and Reintegration Program (RDRP)^5^ | 1998 | 2,822 (2014) | 12,135 | 78% (2000) | <0.5% |
| Rwanda | Genocide Survivors Support and Assistance Fund (FARG)^5^ | 1999 | 21,039 (2013) | 90,468 | 78% (2000) | 1% |
| Rwanda | Vision 2020 Umurenge Programme (VUP)^5^ | 2009 | 246,009 (2015)^4^ | 246,009 | 69.1% (2005) | 3% |
| Sierra Leone | Social Safety Net Programme^26^ | 2015 | 136,768 (2016)^4^ | 136,768 | 54.7% (2011) | 3% |
| South Sudan | Juba Urban Poor Cash Response Pilot^27^ | 2018 | 42,000 (2017) | 42,000 | 44.7% (2009) | 1% |
| Togo | Cash Transfer Programme for Vulnerable Children in Northern Togo^5^ | 2014 | 50,732 (2015)^4^ | 50,732 | 55% | 1% |

### Supplementary Table 3. Characteristics of identified cash transfer programs implemented within included countries prior to the study period (1996 to 2019).

| Country | Program Name | First Complete Year Implemented |
| --- | --- | --- |
| Botswana | Destitute Persons Allowance^5^ | 1981 |
| Eswatini | Public Assistance Grant^5^ | 1987 |
| Mozambique | Programa Subsídio Social Básico^5^ | 1991 |
| Namibia | Multiple programs for elderly, children, disabled^2^ | 1961 |
| South Africa | War Veterans’ Grant^5^ | 1969 |
| Suriname | Social Old Age Pension^28^ | 1974 |
| Thailand | Universal Pension Scheme (for elderly)^5^ | 1994 |
| Zimbabwe | Public Assistance Monthly Maintenance Allowances^5^ | 1988 |

### Supplementary Table 4. Characteristics of females in countries with and without cash transfer programs with total impoverished population coverage greater than 5% included in our individual-level analysis.^*^

|  |  | **Intervention**  **N=355,644** | **Comparison**  **N=939,533** | **Total**  **N=1,295,177** |
| --- | --- | --- | --- | --- |
| **Age, median (IQR)** (N=1,295,177) | | 27 (20-36) | 27 (20-35) | 27 (20-35) |
| **Single marital status** (N=1,287,148) | | 142,508 (40) | 333,916 (36) | 476424 (37) |
| **Education** (N=1,295,136) | **None** | 48,900 (13) | 324,880 (34) | 373,780 (28) |
|  | **Primary** | 163,655 (46) | 331,202 (35) | 494,857 (38) |
|  | **Secondary** | 121,173 (35) | 250,020 (27) | 371,193 (29) |
|  | **Greater than Secondary** | 21,906 (7) | 33,400 (4) | 55,306 (5) |
| **Wealth** (N=1,292,136) | **Richest** | 73,049 (17) | 181,032 (18) | 254,081 (18) |
|  | **Richer** | 66,705 (18) | 174,382 (19) | 241,087 (18) |
|  | **Middle** | 66,515 (19) | 177577 (19) | 244,092 (19) |
|  | **Poorer** | 68,802 (21) | 188657 (21) | 257,459 (21) |
|  | **Poorest** | 80,573 (24) | 214,844 (24) | 295,417 (24) |
| **Rural household** (N=1,295,177) | | 231,404 (64) | 590,207 (63) | 821,611 (63) |
| **PEPFAR funding per capita, median (IQR)** (N=1,295,177) | | 4.4 (0.7-8.0) | 0 (0-2.8) | 0.7 (0-5.9) |
| **HIV Global Fund funding per capita, median (IQR)** (N=1,295,177) | | 1.6 (1.1-4.0) | 0.4 (0-1.2) | 0.8 (0.2-1.6) |
| *World Bank Governance Indicators* | | | |  |
| **Corruption, median (IQR)** (N=1,295,177) | | -0.9 (-1.1 – -0.7) | -0.8 (-1.1 – -0.5) | -0.8 (-1.1 – 0.5) |
| **Stability and Violence, median (IQR)** (N=1,295,177) | | -0.4 (-1.0 – 0.0) | -0.6 (-1.3 – -0.1) | -0.5 (-1.3 – 0.0) |
| **Voice and Accountability, median (IQR)** (N=1,295,177) | | -0.3 (-1.0 – -0.1) | -0.7 (-0.2 – -1.1) | -0.5 (-1.0 - -0.1) |
| **Effectiveness, median (IQR)** (N=1,295,177) | | -0.6 (-0.8 – -0.5) | -0.9 (-1.1 – 0.5) | -0.7 (-1.0 - -0.5) |
| **Rule of Law, median (IQR)** (N=1,295,177) | | -0.4 (-1.0 – -0.3) | -0.9 (-1.1 – -0.5) | -0.7 (-1.1 - -0.4) |
| **Regulatory Quality, median (IQR)** (N=1,295,177) | | -0.5 (-0.8 – -0.3) | -0.7 (-0.9 – -0.4) | -0.5 (-0.9 - -0.3) |
| *Outcomes* | | | |  |
| **Age at sexual debut among youths, median (IQR)** (N=333,091) | | 16 (14-17) | 15 (14-17) | 15 (14-17) |
| **Sexually transmitted infection within 12 months** (N=1,188,596) | | 12,483 (4) | 35,463 (4) | 47,946 (4) |
| **Greater than 1 sexual partner within 12 months** (N=1,117,040) | | 5,926 (2) | 17,637 (2) | 23,563 (2) |
| **HIV test within 2 years** (N=532,037) | | 91,187 (41) | 61,542 (20) | 152,729 (29) |
| **Condom use during last sexual encounter** (N=918,763) | | 31,709 (13) | 56,799 (8) | 88,508 (10) |

^*^N(%) unless otherwise specified

### Supplementary Table 5. Characteristics of males in countries with and without cash transfer programs with total impoverished population coverage greater than 5% included in our individual-level analysis.^*^

|  |  | **Intervention**  **N=190,223** | **Comparison**  **N=400,333** | **Total**  **N=590,556** |
| --- | --- | --- | --- | --- |
| **Age, median (IQR)** (N=590,556) | | 28 (20-39) | 28 (20-39) | 28 (20-39) |
| **Single marital status** (N=589,481) | | 90,238 (47) | 181,072 (45) | 271,310 (46) |
| **Education** (N=590,556) | **None** | 15,693 (8) | 85,245 (21) | 100,938 (17) |
|  | **Primary** | 83,485 (43) | 148,685 (37) | 232,170 (39) |
|  | **Secondary** | 74,617 (40) | 141,232 (35) | 215,849 (37) |
|  | **Greater than Secondary** | 16,418 (9) | 25,132 (7) | 41,550 (7) |
| **Wealth** (N=588,691) | **Richest** | 40,653 (17) | 72,672 (17) | 113,325 (17) |
|  | **Richer** | 36,803 (18) | 72,491 (18) | 109,294 (18) |
|  | **Middle** | 36,312 (20) | 75,128 (19) | 111,440 (19) |
|  | **Poorer** | 36,257 (21) | 81,076 (21) | 117,333 (21) |
|  | **Poorest** | 40,198 (23) | 94,494 (24) | 134,692 (24) |
| **Rural household** (N=590,556) | | 120,886 (61) | 251,866 (63) | 372,752 (62) |
| **PEPFAR funding per capita, median (IQR)** (N=590,556) | | 4.4 (0.6-8.4) | 0 (0-4.4) | 1.5 (0-6.8) |
| **HIV Global Fund funding per capita, median (IQR)** (N=590,556) | | 1.6 (1.0-4.2) | 0.6 (0.2-1.3) | 1.0 (0.3-1.8) |
| *World Bank Governance Indicators* | | | |  |
| **Corruption, median (IQR)** (N=590,556) | | -0.8 (-1.1 - -0.7) | -0.8 (-1.2 - -0.5) | -0.8 (-1.1 - -0.5) |
| **Stability and Violence, median (IQR)** (N=590,556) | | -0.3 (-1.0 - 0.0) | -0.7 (-1.4 - -0.1) | -0.5 (-1.3 - -0.1) |
| **Voice and Accountability, median (IQR)** (N=590,556) | | -0.3 (-1.0 - -0.1) | -0.7 (-1.1 - -0.2) | -0.7 (-1.1 - -0.1) |
| **Effectiveness, median (IQR)** (N=590,556) | | -0.6 (-0.8 - -0.5) | -0.9 (-1.1 - -0.6) | -0.7 (-1.1 - -0.5) |
| **Rule of Law, median (IQR)** (N=590,556) | | -0.6 (-1.0 - -0.3) | -0.9 (-1.2 - -0.5) | -0.7 (-1.1 - -0.4) |
| **Regulatory Quality, median (IQR)** (N=590,556) | | -0.5 (-0.8 - -0.3) | -0.7 (-1.0 - -0.4) | -0.6 (-0.9 - -0.4) |
| *Outcomes* | | | |  |
| **Age at sexual debut among youths, median (IQR)** (N=255,320) | | 15 (14-17) | 16 (14-17) | 16 (14-17) |
| **Sexually transmitted infection within 12 months** (N=564,260) | | 4,763 (3) | 11,867 (3) | 16,630 (3) |
| **Greater than 1 sexual partner within 12 months** (N=549,041) | | 29,026 (16) | 59,455 (17) | 88,481 (16) |
| **HIV test within 2 years** (N=291,323) | | 44,735 (34) | 24,333 (15) | 69,068 (24) |
| **Condom use during last sexual encounter** (N=431,147) | | 35,462 (25) | 55,275 (19) | 90,737 (21) |
| **Transactional sex within 12 months** (N=273,511) | | 5,310 (5) | 6,568 (5) | 12,878 (5) |

^*^N(%) unless otherwise specified

### Supplementary Table 6. Sample description for individual-level outcomes.

| Outcome | Demographic and Health Survey Phases Included | Comparison Observations | Intervention Observations |
| --- | --- | --- | --- |
| *Female* | | | |
| Age at sexual debut among youths | 3,4,5,6,7 | 383,644 | 140,729 |
| Sexually transmitted infection within the prior 12 months | 3,4,5,6,7 | 871,774 | 323,718 |
| Greater than 1 sexual partner within the prior 12 months | 4,5,6,7 | 808,785 | 324,438 |
| HIV test within the prior 12 months | 5,6,7 | 310,682 | 221,355 |
| Condom use during last sexual encounter | 3,4,5,6,7 | 692,149 | 242,398 |
| *Male* | | | |
| Age at sexual debut among youths | 3,4,5,6,7 | 150,723 | 73,162 |
| Sexually transmitted infection within the prior 12 months | 3,4,5,6,7 | 383,181 | 181,079 |
| Greater than 1 sexual partner within the prior 12 months | 4,5,6,7 | 362,704 | 186,337 |
| HIV test within the prior 12 months | 5,6,7 | 158,493 | 132,830 |
| Condom use during last sexual encounter | 3,4,5,6,7 | 290,324 | 140,823 |
| Transactional sex within the prior 12 months | 5,6,7 | 164,439 | 140,823 |

### Supplementary Table 7. The relationship between cash transfer programs and the age of sexual debut among female youths using multivariable linear regression models and including fixed effects for country and year (N=316,064).

|  |  | Coef | 95% Low | 95% High | p-value |
| --- | --- | --- | --- | --- | --- |
| **Cash Program** | | 0.003 | -0.089 | 0.096 | 0.68 |
| Age^1^ |  | 0.283 | 0.254 | 0.311 | <.0001 |
| Single marital status | | 0.191 | 0.052 | 0.331 | 0.009 |
| Education | None | Ref |  |  |  |
|  | Primary | 0.262 | 0.130 | 0.394 | 0.0003 |
|  | Secondary | 1.016 | 0.815 | 1.216 | <.0001 |
|  | Greater than Secondary | 2.193 | 1.902 | 2.484 | <.0001 |
| Wealth | Richest | Ref |  |  |  |
|  | Richer | -0.152 | -0.217 | -0.086 | <.0001 |
|  | Middle | -0.238 | -0.320 | -0.156 | <.0001 |
|  | Poorer | -0.292 | -0.393 | -0.190 | <.0001 |
|  | Poorest | -0.362 | -0.488 | -0.237 | <.0001 |
| Rural |  | -0.027 | -0.082 | 0.028 | 0.32 |
| GDP per capita^2^ | | -0.056 | -0.117 | 0.005 | 0.07 |
| PEPFAR funding per capita^3^ | | 0.030 | -0.006 | 0.066 | 0.10 |
| Global Fund funding per capita^3^ | | 0.012 | -0.094 | 0.118 | 0.82 |
| WB Corruption | | -0.032 | -0.202 | 0.139 | 0.71 |
| WB Stability and Violence | | 0.038 | -0.079 | 0.155 | 0.51 |
| WB Voice and Accountability | | -0.015 | -0.218 | 0.189 | 0.89 |

^1^ per 1 year increase

^2^ per $1000 USD increase

^3^ per $5 USD increase

### Supplementary Table 8. The relationship between cash transfer programs and the age of sexual debut among male youths using multivariable linear regression models and including fixed effects for country and year (N=118,986).

|  |  | Coef | 95% Low | 95% High | p-value |
| --- | --- | --- | --- | --- | --- |
| **Cash Program** | | -0.138 | -0.281 | 0.005 | 0.06 |
| Age^1^ |  | 0.395 | 0.346 | 0.445 | <.0001 |
| Single marital status | | -0.101 | -0.310 | 0.109 | 0.33 |
| Education | None | Ref |  |  |  |
|  | Primary | -0.329 | -0.498 | -0.160 | 0.0004 |
|  | Secondary | -0.181 | -0.378 | 0.017 | 0.07 |
|  | Greater than Secondary | 0.015 | -0.179 | 0.209 | 0.87 |
| Wealth | Richest | Ref |  |  |  |
|  | Richer | -0.044 | -0.126 | 0.038 | 0.28 |
|  | Middle | -0.021 | -0.119 | 0.077 | 0.66 |
|  | Poorer | -0.081 | -0.179 | 0.017 | 0.10 |
|  | Poorest | -0.067 | -0.195 | 0.062 | 0.30 |
| Rural |  | -0.017 | -0.095 | 0.061 | 0.66 |
| GDP per capita^2^ | | -0.082 | -0.242 | 0.079 | 0.31 |
| PEPFAR funding per capita^3^ | | 0.038 | -0.036 | 0.111 | 0.30 |
| Global Fund funding per capita^3^ | | -0.023 | -0.162 | 0.116 | 0.74 |
| WB Corruption | | 0.044 | -0.268 | 0.356 | 0.78 |
| WB Stability and Violence | | 0.126 | -0.105 | 0.357 | 0.27 |
| WB Voice and Accountability | | 0.033 | -0.247 | 0.314 | 0.81 |

^1^ per 1 year increase

^2^ per $1000 USD increase

^3^ per $5 USD increase

### Supplementary Table 9. The relationship between cash transfer programs and sexually transmitted infections within the last 12 months among females using multivariable logistic regression models and including fixed effects for country and year (N=1,179,450).

|  |  | Odds Ratio | 95% Low | 95% High | p-value |
| --- | --- | --- | --- | --- | --- |
| Cash Program |  | 0.67 | 0.50 | 0.91 | 0.01 |
| Age^1^ |  | 1.01 | 1.01 | 1.01 | <.0001 |
| Single marital status |  | 0.60 | 0.53 | 0.68 | <.0001 |
| Education | None | Ref |  |  |  |
|  | Primary | 1.24 | 1.16 | 1.34 | <.0001 |
|  | Secondary | 1.25 | 1.15 | 1.35 | <.0001 |
|  | Greater than Secondary | 1.18 | 1.02 | 1.37 | 0.03 |
| Wealth | Richest | Ref |  |  |  |
|  | Richer | 1.07 | 0.98 | 1.16 | 0.08 |
|  | Middle | 1.01 | 0.90 | 1.14 | 0.53 |
|  | Poorer | 0.95 | 0.82 | 1.11 | 0.88 |
|  | Poorest | 0.86 | 0.73 | 1.02 | 0.12 |
| Rural |  | 0.82 | 0.75 | 0.90 | <.0001 |
| GDP per capita^2^ |  | 0.64 | 0.51 | 0.79 | <.0001 |
| PEPFAR funding per capita^3^ |  | 0.98 | 0.80 | 1.19 | 0.25 |
| Global Fund funding per capita^3^ |  | 0.90 | 0.74 | 1.09 | 0.28 |
| WB Corruption |  | 1.51 | 1.05 | 2.17 | 0.02 |
| WB Stability and Violence |  | 1.02 | 0.79 | 1.33 | 0.87 |
| WB Voice and Accountability |  | 0.82 | 0.49 | 1.35 | 0.43 |

^1^ per 1 year increase

^2^ per $1000 USD increase

^3^ per $5 USD increase

### Supplementary Table 10. The relationship between cash transfer programs and sexually transmitted infections within the last 12 months among males using multivariable logistic regression models and including fixed effects for country and year (N=552,538).

|  |  | Odds Ratio | 95% Low | 95% High | p-value |
| --- | --- | --- | --- | --- | --- |
| Cash Program |  | 1.10 | 0.85 | 1.43 | 0.47 |
| Age^1^ |  | 1.00 | 0.99 | 1.00 | 0.20 |
| Single marital status |  | 0.75 | 0.66 | 0.86 | <.0001 |
| Education | None | Ref |  |  |  |
|  | Primary | 1.17 | 1.06 | 1.30 | 0.003 |
|  | Secondary | 1.19 | 1.04 | 1.35 | 0.01 |
|  | Greater than Secondary | 1.04 | 0.86 | 1.26 | 0.68 |
| Wealth | Richest | Ref |  |  |  |
|  | Richer | 1.09 | 0.99 | 1.20 | 0.07 |
|  | Middle | 1.08 | 0.96 | 1.22 | 0.18 |
|  | Poorer | 1.02 | 0.89 | 1.17 | 0.76 |
|  | Poorest | 0.95 | 0.79 | 1.13 | 0.56 |
| Rural |  | 0.90 | 0.83 | 0.98 | 0.02 |
| GDP per capita^2^ |  | 1.01 | 0.85 | 1.20 | 0.94 |
| PEPFAR funding per capita^3^ |  | 1.01 | 0.90 | 1.13 | 0.92 |
| Global Fund funding per capita^3^ |  | 1.02 | 0.80 | 1.31 | 0.86 |
| WB Corruption |  | 0.89 | 0.53 | 1.50 | 0.67 |
| WB Stability and Violence |  | 1.11 | 0.92 | 1.34 | 0.27 |
| WB Voice and Accountability |  | 1.22 | 0.71 | 2.09 | 0.48 |

^1^ per 1 year increase

^2^ per $1000 USD increase

^3^ per $5 USD increase

### Supplementary Table 11. The relationship between cash transfer programs and having greater than 1 sexual partner in the last 12 months among females using multivariable logistic regression models and including fixed effects for country and year (N=1,110,150).

|  |  | Odds Ratio | 95% Low | 95% High | p-value |
| --- | --- | --- | --- | --- | --- |
| Cash Program |  | 1.04 | 0.75 | 1.46 | 0.80 |
| Age^1^ |  | 1.00 | 0.99 | 1.01 | 0.87 |
| Single marital status |  | 2.04 | 1.65 | 2.52 | <.0001 |
| Education | None | Ref |  |  |  |
|  | Primary | 1.48 | 1.24 | 1.75 | <.0001 |
|  | Secondary | 1.37 | 1.07 | 1.74 | 0.01 |
|  | Greater than Secondary | 1.51 | 1.09 | 2.10 | 0.01 |
| Wealth | Richest | Ref |  |  |  |
|  | Richer | 1.12 | 1.05 | 1.19 | 0.0004 |
|  | Middle | 1.03 | 0.94 | 1.14 | 0.51 |
|  | Poorer | 1.03 | 0.91 | 1.16 | 0.66 |
|  | Poorest | 0.97 | 0.81 | 1.18 | 0.78 |
| Rural |  | 0.77 | 0.69 | 0.86 | <.0001 |
| GDP per capita^2^ |  | 0.75 | 0.44 | 1.28 | 0.29 |
| PEPFAR funding per capita^3^ |  | 1.17 | 0.89 | 1.54 | 0.27 |
| Global Fund funding per capita^3^ |  | 0.88 | 0.67 | 1.15 | 0.35 |
| WB Corruption |  | 0.63 | 0.18 | 2.17 | 0.46 |
| WB Stability and Violence |  | 1.05 | 0.70 | 1.56 | 0.82 |
| WB Voice and Accountability |  | 4.50 | 1.42 | 14.30 | 0.01 |

^1^ per 1 year increase

^2^ per $1000 USD increase

^3^ per $5 USD increase

### Supplementary Table 12. The relationship between cash transfer programs and having greater than 1 sexual partner in the last 12 months among males using multivariable logistic regression models and including fixed effects for country and year (N=540,755).

|  |  | Odds Ratio | 95% Low | 95% High | p-value |
| --- | --- | --- | --- | --- | --- |
| Cash Program |  | 1.12 | 0.99 | 1.28 | 0.07 |
| Age^1^ |  | 1.01 | 1.00 | 1.01 | 0.01 |
| Single marital status |  | 0.65 | 0.55 | 0.76 | <.0001 |
| Education | None | Ref |  |  |  |
|  | Primary | 1.11 | 0.99 | 1.24 | 0.07 |
|  | Secondary | 1.23 | 1.07 | 1.43 | 0.005 |
|  | Greater than Secondary | 1.38 | 1.14 | 1.66 | 0.001 |
| Wealth | Richest | Ref |  |  |  |
|  | Richer | 0.93 | 0.87 | 1.00 | 0.06 |
|  | Middle | 0.91 | 0.82 | 1.00 | 0.05 |
|  | Poorer | 0.85 | 0.76 | 0.95 | 0.005 |
|  | Poorest | 0.78 | 0.68 | 0.90 | 0.0004 |
| Rural |  | 1.11 | 1.03 | 1.20 | 0.01 |
| GDP per capita^2^ |  | 0.94 | 0.80 | 1.11 | 0.48 |
| PEPFAR funding per capita^3^ |  | 1.02 | 0.91 | 1.13 | 0.75 |
| Global Fund funding per capita^3^ |  | 1.00 | 0.91 | 1.09 | 0.97 |
| WB Corruption |  | 1.09 | 0.71 | 1.68 | 0.69 |
| WB Stability and Violence |  | 1.06 | 0.89 | 1.26 | 0.53 |
| WB Voice and Accountability |  | 1.39 | 0.87 | 2.23 | 0.16 |

^1^ per 1 year increase

^2^ per $1000 USD increase

^3^ per $5 USD increase

### Supplementary Table 13. The relationship between cash transfer programs and having an HIV test within the last 12 months among females using multivariable logistic regression models and including fixed effects for country and year (N=525,522).

|  |  | Odds Ratio | 95% Low | 95% High | p-value |
| --- | --- | --- | --- | --- | --- |
| Cash Program |  | 2.61 | 1.15 | 5.88 | 0.02 |
| Age^1^ |  | 0.99 | 0.99 | 1.00 | 0.07 |
| Single marital status |  | 0.47 | 0.42 | 0.53 | <.0001 |
| Education | None | Ref |  |  |  |
|  | Primary | 1.58 | 1.33 | 1.87 | <.0001 |
|  | Secondary | 2.15 | 1.76 | 2.61 | <.0001 |
|  | Greater than Secondary | 3.21 | 2.38 | 4.32 | <.0001 |
| Wealth | Richest | Ref |  |  |  |
|  | Richer | 0.94 | 0.88 | 1.02 | 0.14 |
|  | Middle | 0.87 | 0.78 | 0.96 | 0.01 |
|  | Poorer | 0.81 | 0.70 | 0.92 | 0.002 |
|  | Poorest | 0.76 | 0.64 | 0.90 | 0.002 |
| Rural |  | 0.78 | 0.73 | 0.84 | <.0001 |
| GDP per capita^2^ |  | 0.86 | 0.45 | 1.66 | 0.65 |
| PEPFAR funding per capita^3^ |  | 1.14 | 1.01 | 1.30 | 0.04 |
| Global Fund funding per capita^3^ |  | 1.48 | 1.18 | 1.84 | 0.0005 |
| WB Corruption |  | 0.97 | 0.53 | 1.77 | 0.92 |
| WB Stability and Violence |  | 0.81 | 0.56 | 1.17 | 0.26 |
| WB Voice and Accountability |  | 0.40 | 0.17 | 0.98 | 0.05 |

^1^ per 1 year increase

^2^ per $1000 USD increase

^3^ per $5 USD increase

### Supplementary Table 14. The relationship between cash transfer programs and having an HIV test within the last 12 months among males using multivariable logistic regression models and including fixed effects for country and year (N=291,323).

|  |  | Odds Ratio | 95% Low | 95% High | p-value |
| --- | --- | --- | --- | --- | --- |
| Cash Program |  | 3.19 | 2.45 | 4.15 | <.0001 |
| Age^1^ |  | 1.01 | 1.00 | 1.01 | 0.13 |
| Single marital status |  | 0.57 | 0.51 | 0.64 | <.0001 |
| Education | None | Ref |  |  |  |
|  | Primary | 1.69 | 1.46 | 1.95 | <.0001 |
|  | Secondary | 2.65 | 2.25 | 3.13 | <.0001 |
|  | Greater than Secondary | 4.15 | 3.05 | 5.64 | <.0001 |
| Wealth | Richest | Ref |  |  |  |
|  | Richer | 0.87 | 0.80 | 0.95 | 0.001 |
|  | Middle | 0.80 | 0.70 | 0.91 | 0.001 |
|  | Poorer | 0.74 | 0.62 | 0.88 | 0.001 |
|  | Poorest | 0.64 | 0.51 | 0.80 | 0.0001 |
| Rural |  | 0.85 | 0.79 | 0.91 | <.0001 |
| GDP per capita^2^ |  | 0.61 | 0.42 | 0.89 | 0.01 |
| PEPFAR funding per capita^3^ |  | 1.22 | 1.12 | 1.32 | <.0001 |
| Global Fund funding per capita^3^ |  | 1.22 | 1.09 | 1.36 | 0.0004 |
| WB Corruption |  | 1.36 | 1.03 | 1.78 | 0.03 |
| WB Stability and Violence |  | 0.81 | 0.69 | 0.95 | 0.01 |
| WB Voice and Accountability |  | 0.37 | 0.25 | 0.54 | <.0001 |

^1^ per 1 year increase

^2^ per $1000 USD increase

^3^ per $5 USD increase

### Supplementary Table 15. The relationship between cash transfer programs and condom use during the last sexual encounter among females using multivariable logistic regression models and including fixed effects for country and year (N=918,763).

|  |  | Odds Ratio | 95% Low | 95% High | p-value |
| --- | --- | --- | --- | --- | --- |
| Cash Program |  | 0.94 | 0.77 | 1.14 | 0.50 |
| Age^1^ |  | 0.98 | 0.97 | 0.98 | <.0001 |
| Single marital status |  | 6.56 | 5.54 | 7.77 | <.0001 |
| Education | None | Ref |  |  |  |
|  | Primary | 1.96 | 1.72 | 2.23 | <.0001 |
|  | Secondary | 3.15 | 2.64 | 3.76 | <.0001 |
|  | Greater than Secondary | 3.83 | 3.08 | 4.77 | <.0001 |
| Wealth | Richest | Ref |  |  |  |
|  | Richer | 0.82 | 0.76 | 0.89 | <.0001 |
|  | Middle | 0.69 | 0.62 | 0.76 | <.0001 |
|  | Poorer | 0.61 | 0.55 | 0.68 | <.0001 |
|  | Poorest | 0.50 | 0.44 | 0.56 | <.0001 |
| Rural |  | 0.78 | 0.74 | 0.82 | <.0001 |
| GDP per capita^2^ |  | 1.14 | 1.04 | 1.24 | 0.006 |
| PEPFAR funding per capita^3^ |  | 1.00 | 0.91 | 1.09 | 0.92 |
| Global Fund funding per capita^3^ |  | 1.16 | 0.99 | 1.37 | 0.07 |
| WB Corruption |  | 1.25 | 0.83 | 1.89 | 0.28 |
| WB Stability and Violence |  | 0.90 | 0.71 | 1.15 | 0.42 |
| WB Voice and Accountability |  | 0.87 | 0.67 | 1.12 | 0.27 |

^1^ per 1 year increase

^2^ per $1000 USD increase

^3^ per $5 USD increase

### Supplementary Table 16. The relationship between cash transfer programs and condom use during the last sexual encounter among males using multivariable logistic regression models and including fixed effects for country and year (N=431,147).

|  |  | Odds Ratio | 95% Low | 95% High | p-value |
| --- | --- | --- | --- | --- | --- |
| Cash Program |  | 0.88 | 0.75 | 1.04 | 0.13 |
| Age^1^ |  | 0.98 | 0.97 | 0.98 | <.0001 |
| Single marital status |  | 8.20 | 5.97 | 11.25 | <.0001 |
| Education | None | Ref |  |  |  |
|  | Primary | 1.55 | 1.36 | 1.77 | <.0001 |
|  | Secondary | 2.32 | 1.98 | 2.70 | <.0001 |
|  | Greater than Secondary | 2.72 | 2.25 | 3.28 | <.0001 |
| Wealth | Richest | Ref |  |  |  |
|  | Richer | 0.80 | 0.74 | 0.86 | <.0001 |
|  | Middle | 0.71 | 0.64 | 0.78 | <.0001 |
|  | Poorer | 0.62 | 0.55 | 0.71 | <.0001 |
|  | Poorest | 0.53 | 0.44 | 0.63 | <.0001 |
| Rural |  | 0.80 | 0.74 | 0.86 | <.0001 |
| GDP per capita^2^ |  | 1.00 | 0.86 | 1.15 | 0.95 |
| PEPFAR funding per capita^3^ |  | 1.02 | 0.96 | 1.07 | 0.58 |
| Global Fund funding per capita^3^ |  | 1.00 | 0.89 | 1.14 | 0.95 |
| WB Corruption |  | 1.27 | 1.03 | 1.55 | 0.02 |
| WB Stability and Violence |  | 1.07 | 0.92 | 1.24 | 0.36 |
| WB Voice and Accountability |  | 1.08 | 0.78 | 1.48 | 0.65 |

^1^ per 1 year increase

^2^ per $1000 USD increase

^3^ per $5 USD increase

### Supplementary Table 17. The relationship between cash transfer programs and having transactional sex in the last 12 months among males using multivariable logistic regression models and including fixed effects for country and year (N=273,501).

|  |  | Odds Ratio | 95% Low | 95% High | p-value |
| --- | --- | --- | --- | --- | --- |
| Cash Program |  | 0.99 | 0.85 | 1.15 | 0.86 |
| Age^1^ |  | 0.99 | 0.98 | 1.00 | 0.0008 |
| Single marital status |  | 2.53 | 2.07 | 3.10 | <.0001 |
| Education | None | Ref |  |  |  |
|  | Primary | 1.28 | 1.18 | 1.38 | <.0001 |
|  | Secondary | 1.01 | 0.91 | 1.12 | 0.88 |
|  | Greater than Secondary | 0.76 | 0.66 | 0.88 | 0.0002 |
| Wealth | Richest | Ref |  |  |  |
|  | Richer | 1.17 | 1.01 | 1.34 | 0.04 |
|  | Middle | 1.20 | 0.97 | 1.48 | 0.10 |
|  | Poorer | 1.16 | 0.93 | 1.46 | 0.19 |
|  | Poorest | 1.10 | 0.89 | 1.35 | 0.39 |
| Rural |  | 0.85 | 0.77 | 0.95 | 0.003 |
| GDP per capita^2^ |  | 0.82 | 0.49 | 1.36 | 0.44 |
| PEPFAR funding per capita^3^ |  | 0.93 | 0.80 | 1.09 | 0.39 |
| Global Fund funding per capita^3^ |  | 1.07 | 0.88 | 1.31 | 0.48 |
| WB Corruption |  | 0.74 | 0.38 | 1.41 | 0.36 |
| WB Stability and Violence |  | 1.15 | 0.93 | 1.43 | 0.20 |
| WB Voice and Accountability |  | 1.21 | 0.61 | 2.40 | 0.58 |

^1^ per 1 year increase

^2^ per $1000 USD increase

^3^ per $5 USD increase

### Supplementary Table 18. The relationship between cash transfer programs and number of new HIV infections using multivariable negative binomial models and including fixed effects for country and year.

|  | IRR | 95% low | 95% high | p-value |
| --- | --- | --- | --- | --- |
| **Cash program** | 0.94 | 0.89 | 0.99 | 0.03 |
| GDP per capita ($1000) | 1.00 | 1.00 | 1.01 | 0.41 |
| PEPFAR funding per capita ($5) | 1.00 | 0.99 | 1.01 | 0.63 |
| Global Fund funding per capita ($5) | 0.99 | 0.98 | 1.00 | 0.19 |
| WB Corruption | 1.00 | 0.97 | 1.02 | 0.81 |
| WB Stability and Violence | 1.00 | 0.98 | 1.02 | 0.78 |
| WB Voice and Accountability | 1.02 | 0.98 | 1.05 | 0.36 |

Observations: 985 country-years

### Supplementary Table 19. The relationship between cash transfer programs and number of AIDS-related deaths using multivariable negative binomial models and including fixed effects for country and year.

|  | IRR | 95% low | 95% high | p-value |
| --- | --- | --- | --- | --- |
| **Cash program** | 0.99 | 0.95 | 1.03 | 0.53 |
| GDP per capita ($1000) | 1.00 | 1.00 | 1.01 | 0.42 |
| PEPFAR funding per capita ($5) | 0.98 | 0.97 | 0.99 | 0.004 |
| Global Fund funding per capita ($5) | 0.99 | 0.98 | 1.01 | 0.33 |
| WB Corruption | 0.99 | 0.98 | 1.01 | 0.31 |
| WB Stability and Violence | 1.02 | 0.98 | 1.06 | 0.70 |
| WB Voice and Accountability | 0.99 | 0.97 | 1.02 | 0.59 |

Observations: 985 country-years

### Supplementary Table 20. The relationship between cash transfer programs and the proportion of people with HIV receiving antiretrovirals using multivariable linear regression models and including fixed effects for country and year.

|  | Coef | 95% low | 95% high | p-value |
| --- | --- | --- | --- | --- |
| **Cash program** | 5.0 | -0.2 | 10.1 | 0.06 |
| GDP per capita ($1000) | 2.6 | 1.7 | 3.5 | 0.45 |
| PEPFAR funding per capita ($5) | 3.3 | 0.4 | 6.2 | <.0001 |
| Global Fund funding per capita ($5) | 0.2 | -0.4 | 0.9 | 0.03 |
| WB Corruption | 3.0 | -6.5 | 12.4 | 0.45 |
| WB Stability and Violence | -0.7 | -4.7 | 3.3 | 0.80 |
| WB Voice and Accountability | -4.8 | -11.8 | 2.2 | 0.18 |

Observations: 801 country-years

### Supplementary Table 21. Analysis of interactions for continuous outcomes between cash transfer programs and baseline prevalence at the start of the cash transfer period greater than the median (3.7%) and impoverished population coverage greater than the median (23%).

| Outcome | Subgroup | Coefficient | 95% CI | p-value for interaction |
| --- | --- | --- | --- | --- |
| *Females* | | | | |
| Age at sexual debut among youths | High prevalence | -0.02 | -0.16-0.12 | 0.71 |
|  | Low prevalence | 0.03 | -0.12-0.18 |  |
|  | High coverage | -0.09 | -0.29-0.11 | 0.32 |
|  | Low coverage | 0.03 | -0.07-0.12 |  |
| *Males* | | | | |
| Age at sexual debut among youths | High prevalence | -0.07 | -0.27-0.14 | 0.67 |
|  | Low prevalence | -0.21 | -0.35- -0.07 |  |
|  | High coverage | -0.07 | -0.28-0.14 | 0.82 |
|  | Low coverage | -0.21 | -0.35- -0.07 |  |
| *Country-level* | | | | |
| Proportion of people with HIV receiving ART | High prevalence | 9.7 | 6.1-13.2 | 0.03 |
|  | Low prevalence | 1.0 | -6.7-8.6 |  |
|  | High coverage | 1.6 | -5.2-8.4 | 0.22 |
|  | Low coverage | 7.0 | 0.3-13.7 |  |

### Supplementary Table 22. The relationship between cash transfer programs and the PEPFAR funding per capita (per $5 increase) using multivariable linear regression models and including fixed effects for country and year.

|  | Coef | 95% low | 95% high | p-value |
| --- | --- | --- | --- | --- |
| **Cash program** | **0.3** | **-0.3** | **0.9** | **0.34** |
| GDP per capita ($1000) | 0.03 | -0.06 | 0.12 | 0.53 |
| Global Fund funding per capita ($5) | 0.7 | -0.1 | 1.4 | 0.33 |
| WB Corruption | 0.2 | -0.3 | 0.8 | 0.4 |
| WB Stability and Violence | -0.1 | -0.4 | 0.2 | 0.65 |
| WB Voice and Accountability | -0.1 | -0.6 | 0.4 | 0.75 |

Observations: 985 country-years

### Supplementary Table 23. The relationship between cash transfer programs and HIV-related Global Fund disbursements per capita (per $5 increase) using multivariable linear regression models and including fixed effects for country and year.

|  | Coef | 95% low | 95% high | p-value |
| --- | --- | --- | --- | --- |
| **Cash program** | **0.2** | **-0.1** | **0.5** | **0.08** |
| GDP per capita ($1000) | -0.01 | -0.02 | 0.01 | 0.29 |
| PEPFAR funding per capita ($5) | 0.1 | 0 | 0.18 | 0.06 |
| WB Corruption | 0.1 | -0.1 | 0.3 | 0.49 |
| WB Stability and Violence | 0.1 | 0 | 0.2 | 0.1 |
| WB Voice and Accountability | 0 | -0.2 | 0.1 | 0.51 |

Observations: 985 country-years

### Supplementary Table 24. Primary analysis of the country-level outcome of new HIV infections with exclusion of individual countries to assess for possible outlier countries.

| Country left out | IRR | 95% low | 95% high | p-value |
| --- | --- | --- | --- | --- |
| None | 0.94 | 0.89 | 0.99 | 0.03 |
| Benin | 0.94 | 0.89 | 0.99 | 0.03 |
| Botswana | 0.94 | 0.88 | 0.99 | 0.03 |
| Burkina Faso | 0.94 | 0.88 | 0.99 | 0.03 |
| Burundi | 0.94 | 0.89 | 0.99 | 0.02 |
| Cambodia | 0.94 | 0.89 | 1.00 | 0.04 |
| Cameroon | 0.94 | 0.88 | 0.99 | 0.03 |
| Central African Republic | 0.94 | 0.89 | 0.99 | 0.03 |
| Chad | 0.94 | 0.89 | 0.99 | 0.02 |
| Congo | 0.93 | 0.88 | 0.99 | 0.02 |
| Congo, Dem. Rep. | 0.94 | 0.89 | 0.99 | 0.02 |
| Côte d’Ivoire | 0.94 | 0.89 | 0.99 | 0.02 |
| Djibouti | 0.96 | 0.93 | 1.00 | 0.03 |
| Dominican Republic | 0.94 | 0.89 | 1.00 | 0.04 |
| Equatorial Guinea | 0.94 | 0.89 | 1.00 | 0.03 |
| Eritrea | 0.94 | 0.89 | 0.99 | 0.02 |
| Eswatini | 0.94 | 0.88 | 0.99 | 0.03 |
| Ethiopia | 0.94 | 0.88 | 0.99 | 0.03 |
| Gabon | 0.94 | 0.89 | 0.99 | 0.03 |
| Gambia | 0.94 | 0.89 | 1.00 | 0.03 |
| Ghana | 0.94 | 0.88 | 0.99 | 0.03 |
| Guinea | 0.94 | 0.89 | 0.99 | 0.03 |
| Guinea-Bissau | 0.94 | 0.89 | 0.99 | 0.03 |
| Haiti | 0.94 | 0.88 | 0.99 | 0.03 |
| Jamaica | 0.94 | 0.89 | 1.00 | 0.05 |
| Kenya | 0.94 | 0.88 | 0.99 | 0.03 |
| Lesotho | 0.93 | 0.88 | 0.99 | 0.02 |
| Liberia | 0.94 | 0.89 | 0.99 | 0.02 |
| Malawi | 0.94 | 0.88 | 0.99 | 0.03 |
| Mozambique | 0.94 | 0.89 | 1.00 | 0.03 |
| Namibia | 0.94 | 0.89 | 0.99 | 0.03 |
| Nigeria | 0.94 | 0.89 | 0.99 | 0.03 |
| Rwanda | 0.94 | 0.89 | 0.99 | 0.02 |
| Sierra Leone | 0.94 | 0.89 | 0.99 | 0.03 |
| South Africa | 0.93 | 0.88 | 0.99 | 0.01 |
| South Sudan | 0.94 | 0.89 | 0.99 | 0.03 |
| Suriname | 0.94 | 0.89 | 1.00 | 0.03 |
| Tanzania | 0.93 | 0.88 | 0.99 | 0.02 |
| Thailand | 0.95 | 0.89 | 1.00 | 0.05 |
| Togo | 0.94 | 0.89 | 0.99 | 0.03 |
| Uganda | 0.94 | 0.88 | 0.99 | 0.03 |
| Zambia | 0.93 | 0.88 | 0.99 | 0.02 |
| Zimbabwe | 0.94 | 0.88 | 1.00 | 0.03 |

### Supplementary Table 25. Primary analysis of the individual-level outcome of sexually transmitted infections within the last 12 months for females with exclusion of individual countries to assess for possible outlier countries.

| Country left out | OR | 95% low | 95% high | p-value |
| --- | --- | --- | --- | --- |
| None | 0.67 | 0.50 | 0.91 | 0.01 |
| Benin | 0.71 | 0.52 | 0.98 | 0.04 |
| Botswana | 0.67 | 0.50 | 0.91 | 0.01 |
| Burkina Faso | 0.68 | 0.50 | 0.93 | 0.01 |
| Burundi | 0.67 | 0.49 | 0.91 | 0.01 |
| Cambodia | 0.66 | 0.48 | 0.92 | 0.01 |
| Cameroon | 0.55 | 0.44 | 0.69 | <.0001 |
| Central African Republic | 0.67 | 0.50 | 0.91 | 0.01 |
| Chad | 0.67 | 0.49 | 0.92 | 0.01 |
| Congo | 0.67 | 0.49 | 0.91 | 0.01 |
| Congo, Dem. Rep. | 0.67 | 0.49 | 0.91 | 0.01 |
| Côte d’Ivoire | 0.66 | 0.48 | 0.89 | 0.008 |
| Djibouti | 0.67 | 0.50 | 0.91 | 0.01 |
| Dominican Republic | 0.77 | 0.58 | 1.02 | 0.07 |
| Equatorial Guinea | 0.67 | 0.50 | 0.91 | 0.01 |
| Eritrea | 0.67 | 0.50 | 0.91 | 0.01 |
| Eswatini | 0.68 | 0.50 | 0.92 | 0.01 |
| Ethiopia | 0.68 | 0.50 | 0.93 | 0.01 |
| Gabon | 0.67 | 0.49 | 0.90 | 0.009 |
| Gambia | 0.67 | 0.50 | 0.91 | 0.01 |
| Ghana | 0.67 | 0.48 | 0.92 | 0.01 |
| Guinea | 0.66 | 0.50 | 0.88 | 0.005 |
| Guinea-Bissau | 0.67 | 0.50 | 0.91 | 0.01 |
| Haiti | 0.68 | 0.49 | 0.96 | 0.03 |
| Jamaica | 0.67 | 0.50 | 0.91 | 0.01 |
| Kenya | 0.68 | 0.50 | 0.93 | 0.02 |
| Lesotho | 0.67 | 0.49 | 0.90 | 0.008 |
| Liberia | 0.68 | 0.50 | 0.93 | 0.02 |
| Malawi | 0.67 | 0.49 | 0.91 | 0.01 |
| Mozambique | 0.67 | 0.48 | 0.93 | 0.02 |
| Namibia | 0.68 | 0.50 | 0.91 | 0.01 |
| Nigeria | 0.73 | 0.53 | 1.00 | 0.05 |
| Rwanda | 0.66 | 0.48 | 0.91 | 0.01 |
| Sierra Leone | 0.69 | 0.51 | 0.92 | 0.01 |
| South Africa | 0.67 | 0.50 | 0.91 | 0.01 |
| South Sudan | 0.67 | 0.50 | 0.91 | 0.01 |
| Suriname | 0.67 | 0.50 | 0.91 | 0.01 |
| Tanzania | 0.73 | 0.55 | 0.96 | 0.03 |
| Thailand | 0.67 | 0.50 | 0.91 | 0.01 |
| Togo | 0.69 | 0.50 | 0.95 | 0.02 |
| Uganda | 0.57 | 0.42 | 0.78 | 0.0004 |
| Zambia | 0.71 | 0.54 | 0.93 | 0.01 |
| Zimbabwe | 0.66 | 0.48 | 0.92 | 0.01 |

### Supplementary Table 26. Primary analysis of the individual-level outcome of HIV testing within the last 12 months for females with exclusion of individual countries to assess for possible outlier countries.

| Country left out | OR | 95% low | 95% high | p-value |
| --- | --- | --- | --- | --- |
| None | 2.61 | 1.15 | 5.89 | 0.02 |
| Benin | 1.51 | 1.06 | 2.14 | 0.02 |
| Botswana | 2.61 | 1.15 | 5.88 | 0.02 |
| Burkina Faso | 2.61 | 1.15 | 5.88 | 0.02 |
| Burundi | 2.67 | 1.18 | 6.05 | 0.02 |
| Cambodia | 1.72 | 1.03 | 2.34 | 0.04 |
| Cameroon | 2.75 | 1.17 | 6.47 | 0.02 |
| Central African Republic | 2.61 | 1.15 | 5.89 | 0.02 |
| Chad | 2.59 | 1.15 | 5.88 | 0.02 |
| Congo | 2.70 | 1.14 | 6.35 | 0.02 |
| Congo, Dem. Rep. | 2.60 | 1.15 | 5.88 | 0.02 |
| Côte d’Ivoire | 2.61 | 1.15 | 5.88 | 0.02 |
| Djibouti | 2.61 | 1.15 | 5.88 | 0.02 |
| Dominican Republic | 2.61 | 1.15 | 5.92 | 0.02 |
| Equatorial Guinea | 2.61 | 1.15 | 5.88 | 0.02 |
| Eritrea | 2.61 | 1.15 | 5.88 | 0.02 |
| Eswatini | 2.57 | 1.15 | 5.77 | 0.02 |
| Ethiopia | 2.60 | 1.15 | 5.86 | 0.02 |
| Gabon | 2.61 | 1.15 | 5.88 | 0.02 |
| Gambia | 2.61 | 1.15 | 5.88 | 0.02 |
| Ghana | 2.61 | 1.15 | 5.88 | 0.02 |
| Guinea | 6.02 | 3.65 | 9.94 | <.0001 |
| Guinea-Bissau | 2.61 | 1.15 | 5.88 | 0.02 |
| Haiti | 3.48 | 2.00 | 6.06 | <.0001 |
| Jamaica | 2.61 | 1.15 | 5.88 | 0.02 |
| Kenya | 2.44 | 1.06 | 5.64 | 0.04 |
| Lesotho | 2.55 | 1.13 | 5.72 | 0.02 |
| Liberia | 2.60 | 1.15 | 5.88 | 0.02 |
| Malawi | 2.32 | 0.91 | 5.90 | 0.08 |
| Mozambique | 2.29 | 0.78 | 6.70 | 0.13 |
| Namibia | 2.41 | 1.02 | 5.66 | 0.04 |
| Nigeria | 2.60 | 1.15 | 5.85 | 0.02 |
| Rwanda | 2.65 | 1.44 | 4.88 | 0.002 |
| Sierra Leone | 2.87 | 1.18 | 7.02 | 0.02 |
| South Africa | 2.61 | 1.16 | 5.90 | 0.02 |
| South Sudan | 2.61 | 1.15 | 5.88 | 0.02 |
| Suriname | 2.61 | 1.15 | 5.88 | 0.02 |
| Tanzania | 2.54 | 1.11 | 5.94 | 0.03 |
| Thailand | 2.61 | 1.15 | 5.88 | 0.02 |
| Togo | 2.61 | 1.15 | 5.88 | 0.02 |
| Uganda | 2.49 | 1.18 | 5.24 | 0.02 |
| Zambia | 1.44 | 0.48 | 4.38 | 0.52 |
| Zimbabwe | 2.85 | 1.42 | 5.73 | 0.003 |

### Supplementary Table 27. Primary analysis of the individual-level outcome of HIV testing within the last 12 months for males with exclusion of individual countries to assess for possible outlier countries.

| Country left out | OR | 95% low | 95% high | p-value |
| --- | --- | --- | --- | --- |
| None | 3.19 | 2.45 | 4.15 | <.0001 |
| Benin | 2.91 | 2.43 | 3.48 | <.0001 |
| Botswana | 3.19 | 2.45 | 4.15 | <.0001 |
| Burkina Faso | 3.19 | 2.45 | 4.15 | <.0001 |
| Burundi | 3.15 | 2.43 | 4.07 | <.0001 |
| Cambodia | 2.91 | 2.56 | 3.31 | <.0001 |
| Cameroon | 3.34 | 2.65 | 4.21 | <.0001 |
| Central African Republic | 3.19 | 2.45 | 4.15 | <.0001 |
| Chad | 3.11 | 2.40 | 4.05 | <.0001 |
| Congo | 3.40 | 2.64 | 4.38 | <.0001 |
| Congo, Dem. Rep. | 3.19 | 2.45 | 4.15 | <.0001 |
| Côte d’Ivoire | 3.19 | 2.45 | 4.15 | <.0001 |
| Djibouti | 3.19 | 2.45 | 4.15 | <.0001 |
| Dominican Republic | 3.21 | 2.46 | 4.20 | <.0001 |
| Equatorial Guinea | 3.19 | 2.45 | 4.15 | <.0001 |
| Eritrea | 3.19 | 2.45 | 4.15 | <.0001 |
| Eswatini | 3.19 | 2.43 | 4.19 | <.0001 |
| Ethiopia | 3.18 | 2.45 | 4.13 | <.0001 |
| Gabon | 3.19 | 2.45 | 4.15 | <.0001 |
| Gambia | 3.19 | 2.45 | 4.15 | <.0001 |
| Ghana | 3.19 | 2.45 | 4.15 | <.0001 |
| Guinea | 3.43 | 2.66 | 4.23 | <.0001 |
| Guinea-Bissau | 3.19 | 2.45 | 4.15 | <.0001 |
| Haiti | 3.98 | 3.05 | 5.19 | <.0001 |
| Jamaica | 3.19 | 2.45 | 4.15 | <.0001 |
| Kenya | 3.23 | 2.49 | 4.20 | <.0001 |
| Lesotho | 3.14 | 2.32 | 4.25 | <.0001 |
| Liberia | 3.18 | 2.44 | 4.14 | <.0001 |
| Malawi | 2.69 | 1.79 | 4.03 | <.0001 |
| Mozambique | 3.19 | 2.45 | 4.16 | <.0001 |
| Namibia | 3.09 | 2.31 | 4.13 | <.0001 |
| Nigeria | 3.19 | 2.45 | 4.14 | <.0001 |
| Rwanda | 3.48 | 2.82 | 4.31 | <.0001 |
| Sierra Leone | 3.22 | 2.48 | 4.18 | <.0001 |
| South Africa | 3.19 | 2.45 | 4.16 | <.0001 |
| South Sudan | 3.19 | 2.45 | 4.15 | <.0001 |
| Suriname | 3.19 | 2.45 | 4.15 | <.0001 |
| Tanzania | 2.06 | 1.50 | 2.83 | <.0001 |
| Thailand | 3.19 | 2.45 | 4.15 | <.0001 |
| Togo | 3.19 | 2.45 | 4.15 | <.0001 |
| Uganda | 3.07 | 2.36 | 4.00 | <.0001 |
| Zambia | 2.50 | 1.94 | 3.21 | <.0001 |
| Zimbabwe | 3.47 | 2.42 | 4.98 | <.0001 |

### Supplementary Table 28. Test of parallel trends assumption: risk of sexually transmitted infection within last 12 months, prior to cash transfer programs, among females.

|  |  | Odds Ratio | 95% Low | 95% High | p-value |
| --- | --- | --- | --- | --- | --- |
| Cash Program Country | | 2.33 | 0.87 | 6.26 | 0.09 |
| Time Trend^1^ |  | 0.77 | 0.52 | 1.15 | 0.21 |
| **Cash Program Country * Time Trend** | | **0.98** | **0.92** | **1.04** | **0.47** |
| Age^1^ |  | 1.01 | 1.00 | 1.01 | 0.0001 |
| Single marital status | | 0.63 | 0.56 | 0.70 | <.0001 |
| Education | None | Ref |  |  |  |
|  | Primary | 1.21 | 1.11 | 1.32 | <.0001 |
|  | Secondary | 1.25 | 1.14 | 1.36 | <.0001 |
|  | Greater than Secondary | 1.22 | 1.08 | 1.37 | 0.001 |
| Wealth | Richest | Ref |  |  |  |
|  | Richer | 1.04 | 0.95 | 1.14 | 0.43 |
|  | Middle | 0.98 | 0.85 | 1.12 | 0.74 |
|  | Poorer | 0.92 | 0.79 | 1.08 | 0.32 |
|  | Poorest | 0.82 | 0.72 | 0.93 | 0.003 |
| Rural |  | 0.81 | 0.74 | 0.87 | <.0001 |
| GDP per capita^2^ | | 0.69 | 0.33 | 1.44 | 0.33 |
| PEPFAR funding per capita^3^ | | 1.15 | 0.94 | 1.42 | 0.17 |
| Global Fund funding per capita^3^ | | 0.97 | 0.55 | 1.71 | 0.91 |
| WB Corruption | | 1.43 | 0.77 | 2.68 | 0.26 |
| WB Stability and Violence | | 1.04 | 0.77 | 1.40 | 0.81 |
| WB Voice and Accountability | | 0.89 | 0.36 | 2.21 | 0.80 |

^1^ per increase in 1 year

^2^ per increase in $1000 USD

^3^ per increase in $5 USD

### Supplementary Table 29. Test of parallel trends assumption: risk of sexually transmitted infection within last 12 months, prior to cash transfer programs, among males.

|  |  | Odds Ratio | 95% Low | 95% High | p-value |
| --- | --- | --- | --- | --- | --- |
| Cash Program Country | | 2.38 | 1.41 | 4.03 | 0.001 |
| Time Trend^1^ | | 0.77 | 0.38 | 1.59 | 0.49 |
| **Cash Program Country * Time Trend** | | **0.99** | **0.95** | **1.02** | **0.49** |
| Age^1^ |  | 1.00 | 0.99 | 1.00 | 0.09 |
| Single marital status | | 0.78 | 0.69 | 0.88 | <.0001 |
| Education | None | Ref |  |  |  |
|  | Primary | 1.18 | 1.05 | 1.33 | 0.007 |
|  | Secondary | 1.19 | 1.03 | 1.37 | 0.02 |
|  | Greater than Secondary | 1.05 | 0.86 | 1.29 | 0.64 |
| Wealth | Richest | Ref |  |  |  |
|  | Richer | 1.08 | 0.97 | 1.21 | 0.17 |
|  | Middle | 1.09 | 0.95 | 1.25 | 0.20 |
|  | Poorer | 1.02 | 0.87 | 1.19 | 0.82 |
|  | Poorest | 0.98 | 0.80 | 1.20 | 0.83 |
| Rural |  | 0.89 | 0.82 | 0.98 | 0.02 |
| GDP per capita^2^ | | 0.88 | 0.69 | 1.12 | 0.29 |
| PEPFAR funding per capita^3^ | | 1.23 | 1.11 | 1.37 | 0.0001 |
| Global Fund funding per capita^3^ | | 1.03 | 0.64 | 1.64 | 0.92 |
| WB Corruption | | 1.01 | 0.59 | 1.73 | 0.97 |
| WB Stability and Violence | | 1.03 | 0.82 | 1.30 | 0.80 |
| WB Voice and Accountability | | 1.15 | 0.67 | 1.97 | 0.61 |

^1^ per increase in 1 year

^2^ per increase in $1000 USD

^3^ per increase in $5 USD

### Supplementary Table 30. Test of parallel trends assumption: number of new HIV infections, prior to cash transfer programs.

|  | IRR | 95% Low | 95% High | p-value |
| --- | --- | --- | --- | --- |
| Cash Program Country | 7.08 | 6.17 | 8.12 | <.0001 |
| Time Trend^1^ | 0.94 | 0.91 | 0.98 | 0.001 |
| **Cash Program Country * Time Trend** | **0.99** | **0.96** | **1.02** | **0.55** |
| GDP per capita^2^ | 1.00 | 1.00 | 1.01 | 0.16 |
| PEPFAR funding per capita^3^ | 0.97 | 0.95 | 0.98 | 0.0003 |
| Global Fund funding per capita^3^ | 0.99 | 0.98 | 1.01 | 0.49 |
| WB Corruption | 1.02 | 0.97 | 1.07 | 0.42 |
| WB Stability and Violence | 0.99 | 0.97 | 1.02 | 0.52 |
| WB Voice and Accountability | 1.00 | 0.95 | 1.06 | 0.91 |

^1^ per increase in 1 year

^2^ per increase in $5 USD

^3^ per increase in $1000 USD

### Supplementary Table 31. Test of parallel trends assumption: HIV test within last 12 months, prior to cash transfer programs, among females.

|  |  | Odds Ratio | 95% Low | 95% High | p-value |
| --- | --- | --- | --- | --- | --- |
| Cash Program Country | | 54.60 | 40.33 | 73.93 | <.0001 |
| Time Trend^1^ |  | 2.73 | 2.64 | 2.82 | <.0001 |
| **Cash Program Country * Time Trend** | | 0.81 | 0.81 | 0.82 | <.0001 |
| Age^1^ |  | 0.99 | 0.99 | 1.00 | 0.0006 |
| Single marital status | | 0.48 | 0.41 | 0.56 | <.0001 |
| Education | None | Ref |  |  |  |
|  | Primary | 1.45 | 1.22 | 1.74 | <.0001 |
|  | Secondary | 2.08 | 1.60 | 2.71 | <.0001 |
|  | Greater than Secondary | 4.13 | 2.94 | 5.80 | <.0001 |
| Wealth | Richest | Ref |  |  |  |
|  | Richer | 0.88 | 0.80 | 0.97 | 0.0073 |
|  | Middle | 0.76 | 0.65 | 0.88 | 0.0003 |
|  | Poorer | 0.68 | 0.56 | 0.82 | <.0001 |
|  | Poorest | 0.62 | 0.48 | 0.80 | 0.0003 |
| Rural |  | 0.76 | 0.67 | 0.85 | <.0001 |
| GDP per capita^2^ | | 0.30 | 0.28 | 0.32 | <.0001 |
| PEPFAR funding per capita^3^ | | 0.79 | 0.78 | 0.80 | <.0001 |
| Global Fund funding per capita^3^ | | 0.41 | 0.35 | 0.48 | <.0001 |
| WB Corruption | | 0.05 | 0.05 | 0.06 | <.0001 |
| WB Stability and Violence | | 30.97 | 29.07 | 33.00 | <.0001 |
| WB Voice and Accountability | | 0.72 | 0.63 | 0.83 | <.0001 |

^1^ per increase in 1 year

^2^ per increase in $1000 USD

^3^ per increase in $5 USD

### Supplementary Table 32. Test of parallel trends assumption: HIV test within last 12 months, prior to cash transfer programs, among males.

|  |  | Odds Ratio | 95% Low | 95% High | p-value |
| --- | --- | --- | --- | --- | --- |
| Cash Program Country | | 0.50 | 0.33 | 0.74 | 0.0005 |
| Time Trend^1^ |  | 1.07 | 1.01 | 1.12 | 0.01 |
| **Cash Program Country * Time Trend** | | 1.27 | 1.25 | 1.29 | <.0001 |
| Age^1^ |  | 0.99 | 0.99 | 1.00 | 0.28 |
| Single marital status | | 0.48 | 0.41 | 0.56 | <.0001 |
| Education | None | Ref |  |  |  |
|  | Primary | 1.45 | 1.22 | 1.74 | <.0001 |
|  | Secondary | 2.08 | 1.60 | 2.71 | <.0001 |
|  | Greater than Secondary | 4.13 | 2.94 | 5.80 | <.0001 |
| Wealth | Richest | Ref |  |  |  |
|  | Richer | 0.88 | 0.80 | 0.97 | <.0001 |
|  | Middle | 0.76 | 0.65 | 0.88 | 0.0004 |
|  | Poorer | 0.68 | 0.56 | 0.82 | 0.0002 |
|  | Poorest | 0.62 | 0.48 | 0.80 | 0.001 |
| Rural |  | 0.76 | 0.67 | 0.85 | 0.02 |
| GDP per capita^2^ | | 0.30 | 0.28 | 0.32 | 0.05 |
| PEPFAR funding per capita^3^ | | 0.79 | 0.78 | 0.80 | 0.58 |
| Global Fund funding per capita^3^ | | 0.41 | 0.35 | 0.48 | <.0001 |
| WB Corruption | | 0.05 | 0.05 | 0.06 | <.0001 |
| WB Stability and Violence | | 30.97 | 29.07 | 33.00 | <.0001 |
| WB Voice and Accountability | | 0.72 | 0.63 | 0.83 | 0.0002 |

^1^ per increase in 1 year

^2^ per increase in $1000 USD

^3^ per increase in $5 USD

### Supplementary Table 33. Repeat of the primary analyses with exclusion of country-years after the fourth year of the cash transfer program.

| Outcome | Effect measure | 95% low | 95% high | p-value |
| --- | --- | --- | --- | --- |
| Incidence | 0.94 | 0.89 | 0.99 | 0.02 |
| AIDS-related Deaths | 0.99 | 0.95 | 1.03 | 0.56 |
| ART Coverage | 3.4 | -0.2 | 6.9 | 0.06 |
| Individual-level, Females | | | | |
| Age at sexual debut among youths | 0.03 | -0.08 | 0.14 | 0.59 |
| Sexually transmitted infection within 12 months | 0.78 | 0.60 | 1.01 | 0.06 |
| Greater than 1 sexual partner within 12 months | 0.97 | 0.70 | 1.36 | 0.87 |
| HIV test within 12 months | 5.86 | 0.93 | 36.93 | 0.06 |
| Condom use at last sex | 0.94 | 0.77 | 1.16 | 0.56 |
| Individual-level, Males | | | | |
| Age at sexual debut among youths | -0.14 | -0.3 | 0.02 | 0.09 |
| Sexually transmitted infection within 12 months | 1.11 | 0.87 | 1.43 | 0.4 |
| Greater than 1 sexual partner within 12 months | 1.12 | 0.96 | 1.29 | 0.15 |
| HIV test within 12 months | 3.74 | 2.60 | 5.36 | <.0001 |
| Condom use at last sex | 0.86 | 0.76 | 1.02 | 0.08 |
| Transactional sex within 12 months | 1.12 | 0.87 | 1.44 | 0.37 |

### Supplementary Table 34. Repeat of the primary analyses with exclusion of intervention countries that have negative weights during the early years of the cash transfer program (Jamaica, Ghana, Dominican Republic, Congo).

| Outcome | Effect measure | 95% low | 95% high | p-value |
| --- | --- | --- | --- | --- |
| Incidence | 0.94 | 0.87 | 1.00 | 0.06 |
| AIDS-related Deaths | 0.98 | 0.93 | 1.03 | 0.39 |
| ART Coverage | 7.6 | 2.8 | 12.3 | 0.003 |
| Individual-level, Females | | | | |
| Age at sexual debut among youths | 0.03 | -0.07 | 0.14 | 0.52 |
| Sexually transmitted infection within 12 months | 0.77 | 0.57 | 1.04 | 0.08 |
| Greater than 1 sexual partner within 12 months | 1.16 | 0.89 | 1.52 | 0.28 |
| HIV test within 12 months | 2.70 | 1.14 | 6.40 | 0.02 |
| Condom use at last sex | 0.96 | 0.77 | 1.21 | 0.75 |
| Individual-level, Males | | | | |
| Age at sexual debut among youths | -0.09 | -0.27 | 0.09 | 0.3 |
| Sexually transmitted infection within 12 months | 1.21 | 0.95 | 1.54 | 0.13 |
| Greater than 1 sexual partner within 12 months | 1.09 | 0.92 | 1.29 | 0.31 |
| HIV test within 12 months | 3.43 | 2.65 | 4.44 | <.0001 |
| Condom use at last sex | 0.89 | 0.75 | 1.07 | 0.22 |
| Transactional sex within 12 months | 1.04 | 0.87 | 1.24 | 0.69 |

### Supplementary Figure 1. Trends in annual HIV incidence rate per 100,000 persons by country. The vertical dashed line represents the start of cash transfer program period.

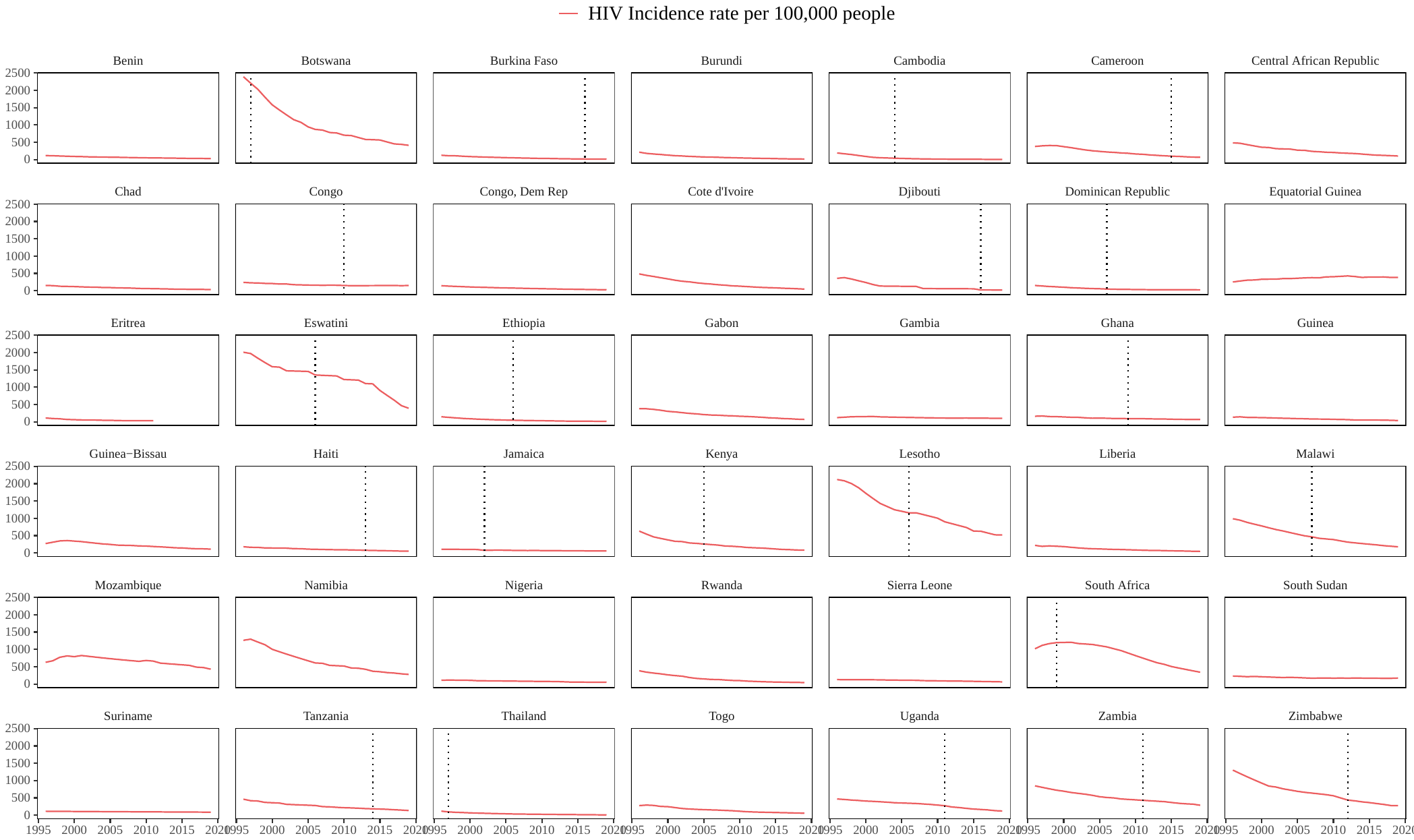

### Supplementary Figure 2. Trends in annual incidence rate of AIDS-related deaths per 100,000 persons by country. The dashed line represents the start of cash transfer program period.

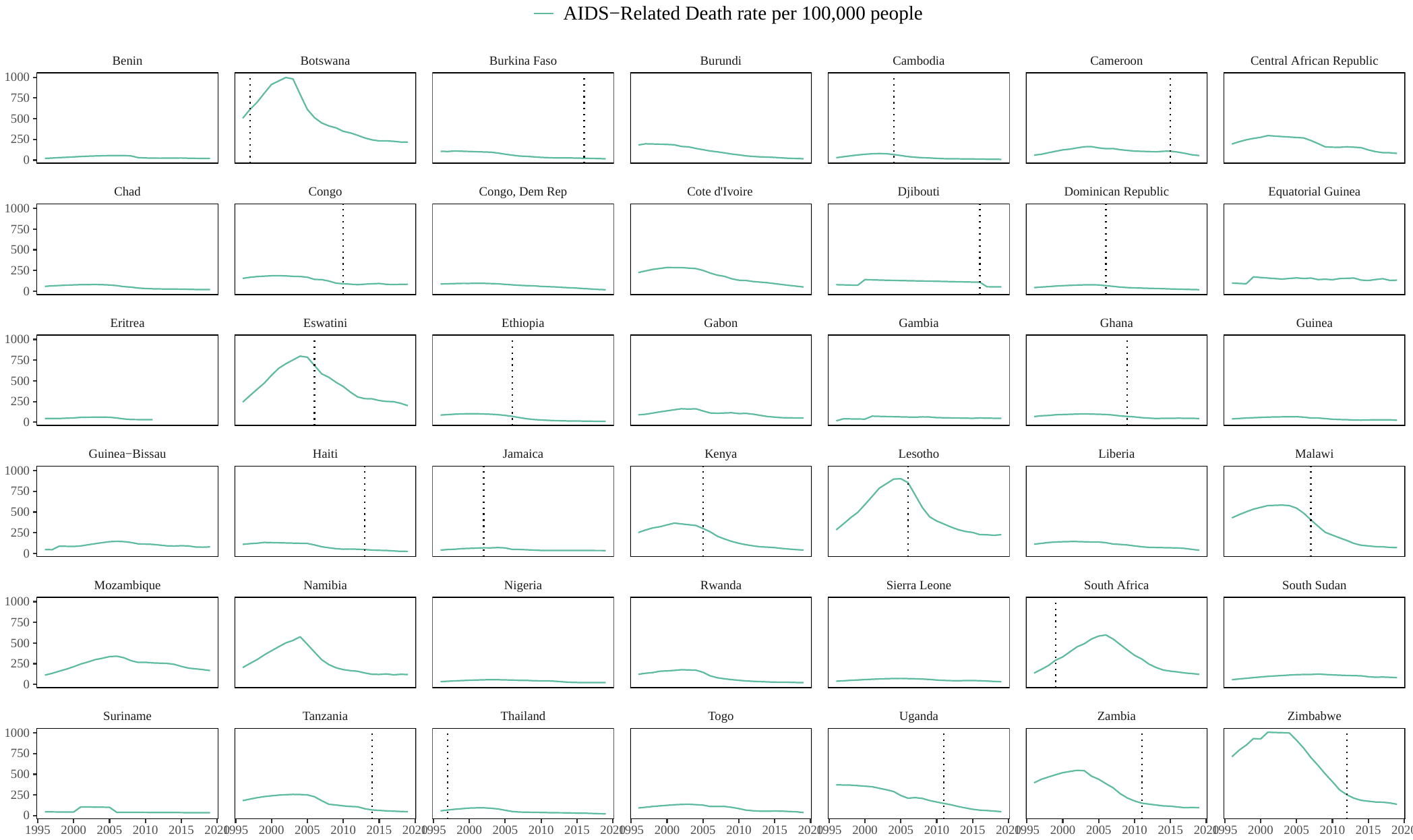

### Supplementary Figure 3. Trends in the proportion of people with HIV receiving antiretroviral therapy by country. The dashed line represents the start of cash transfer program period.

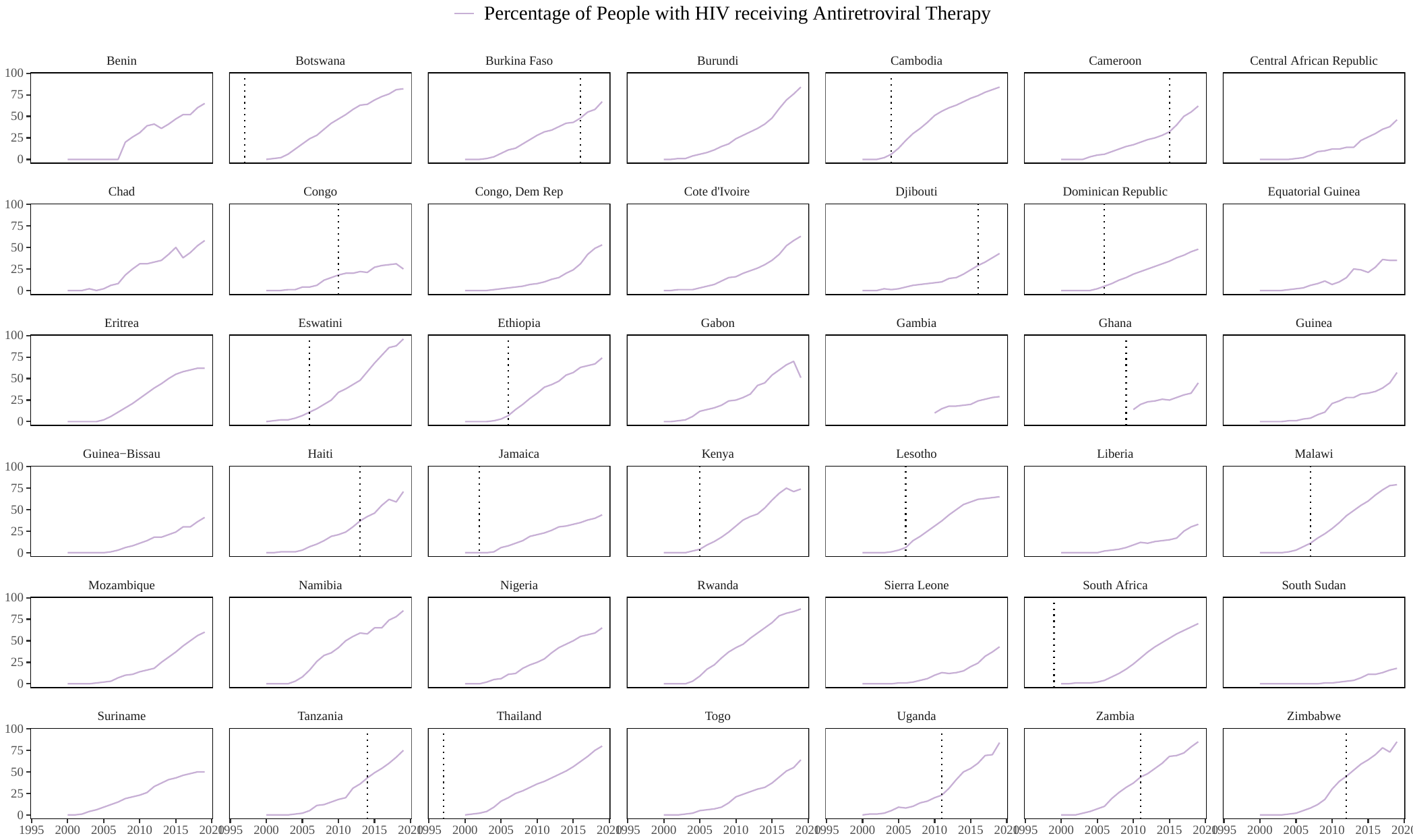

Supplementary Figure 4. Forest plot of individual-level outcomes stratified by wealth quintile. Multivariable models include cash transfer program, age, single marital status, education, rural household setting, and the country-level covariates PEPFAR funding per capita, HIV-related Global Fund disbursements per capita, and three World Bank Governance Indicators: Corruption, Stability and Violence, and Voice and Accountability.
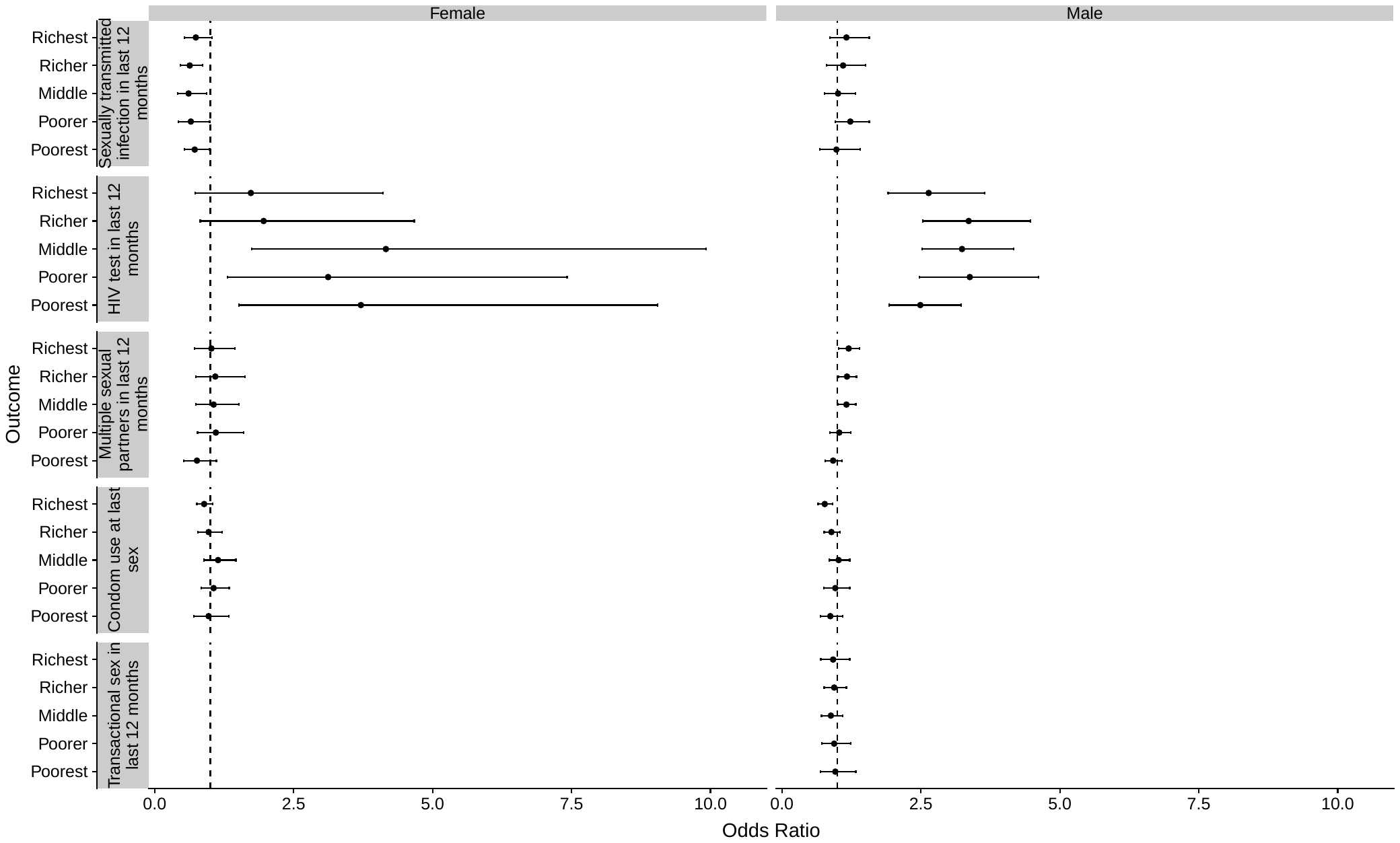

Supplementary Figure 5. Adjusted odds ratios of sexually transmitted infections and HIV testing within the last year among females as a function of year of the cash transfer period, with several years categorized together because of sample size.

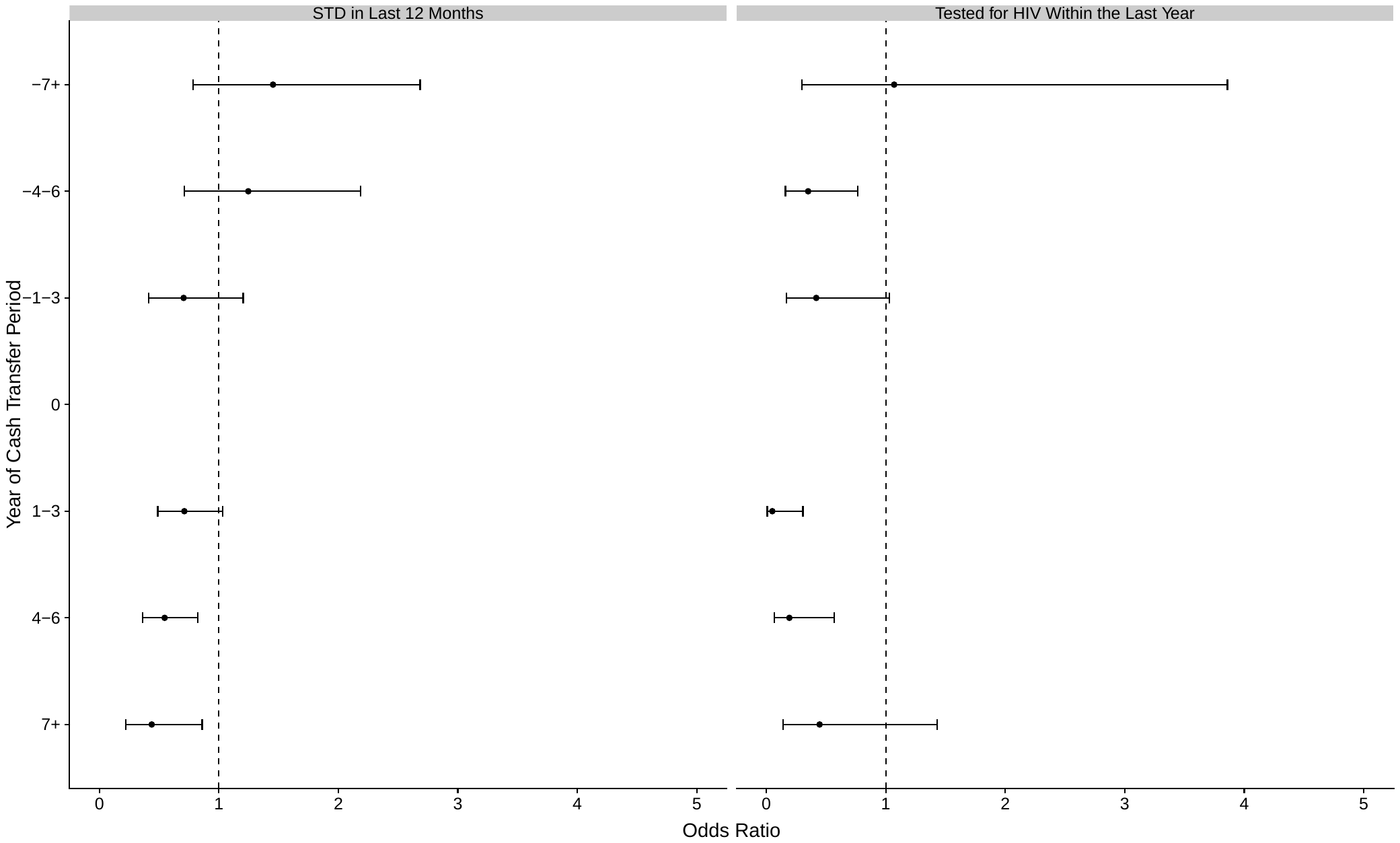

Supplementary Figure 6. Adjusted odds ratios of sexually transmitted infections and HIV testing within the last year among males as a function of year of the cash transfer period, with several years categorized together because of sample size.

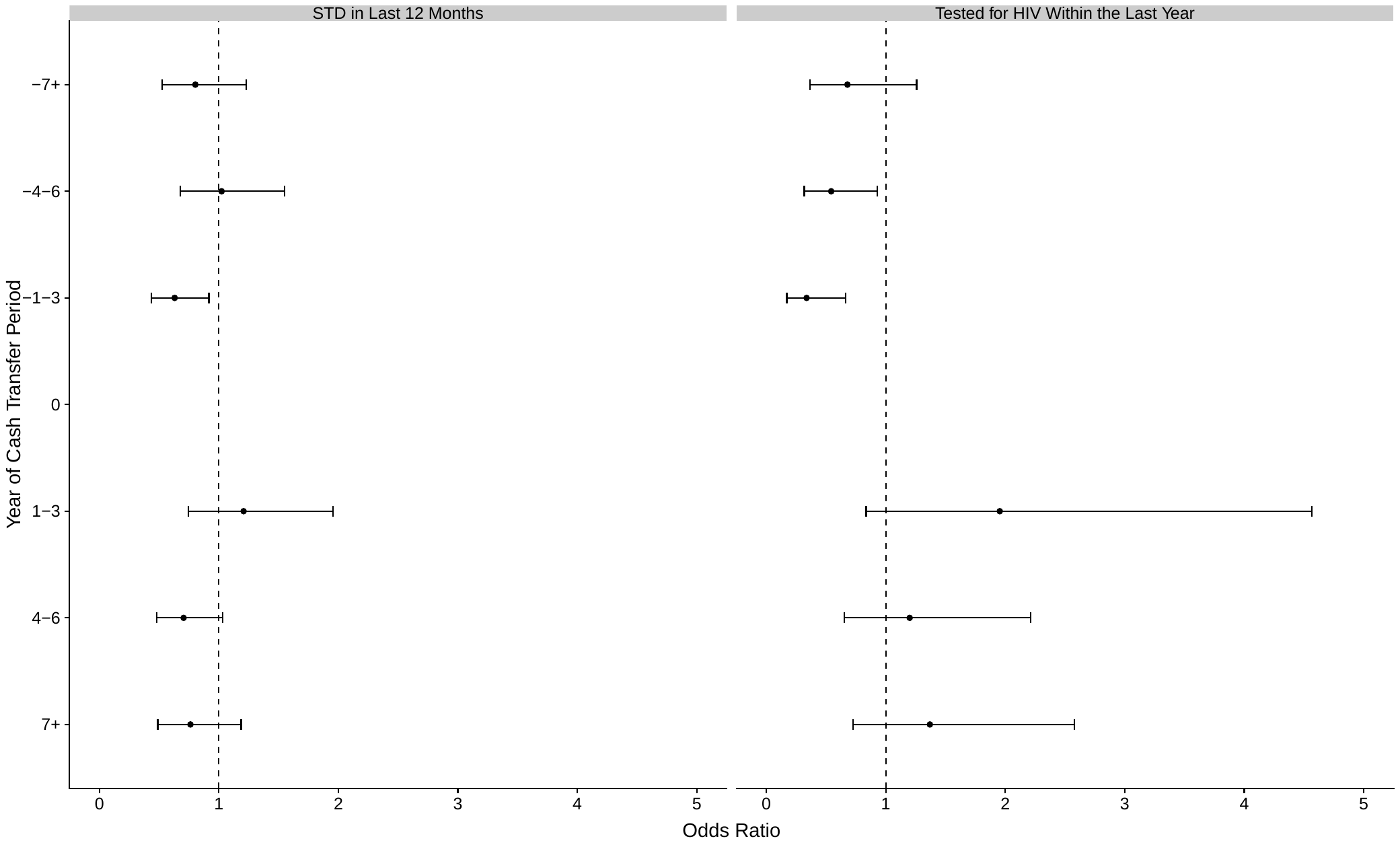

### Supplementary Figure 7. Histogram of residuals from regression of relationship between cash transfer programs and the proportion of people with HIV receiving antiretrovirals using multivariable linear regression models and including fixed effects for country and year, scaled by the sum of the squared residuals across all observations.

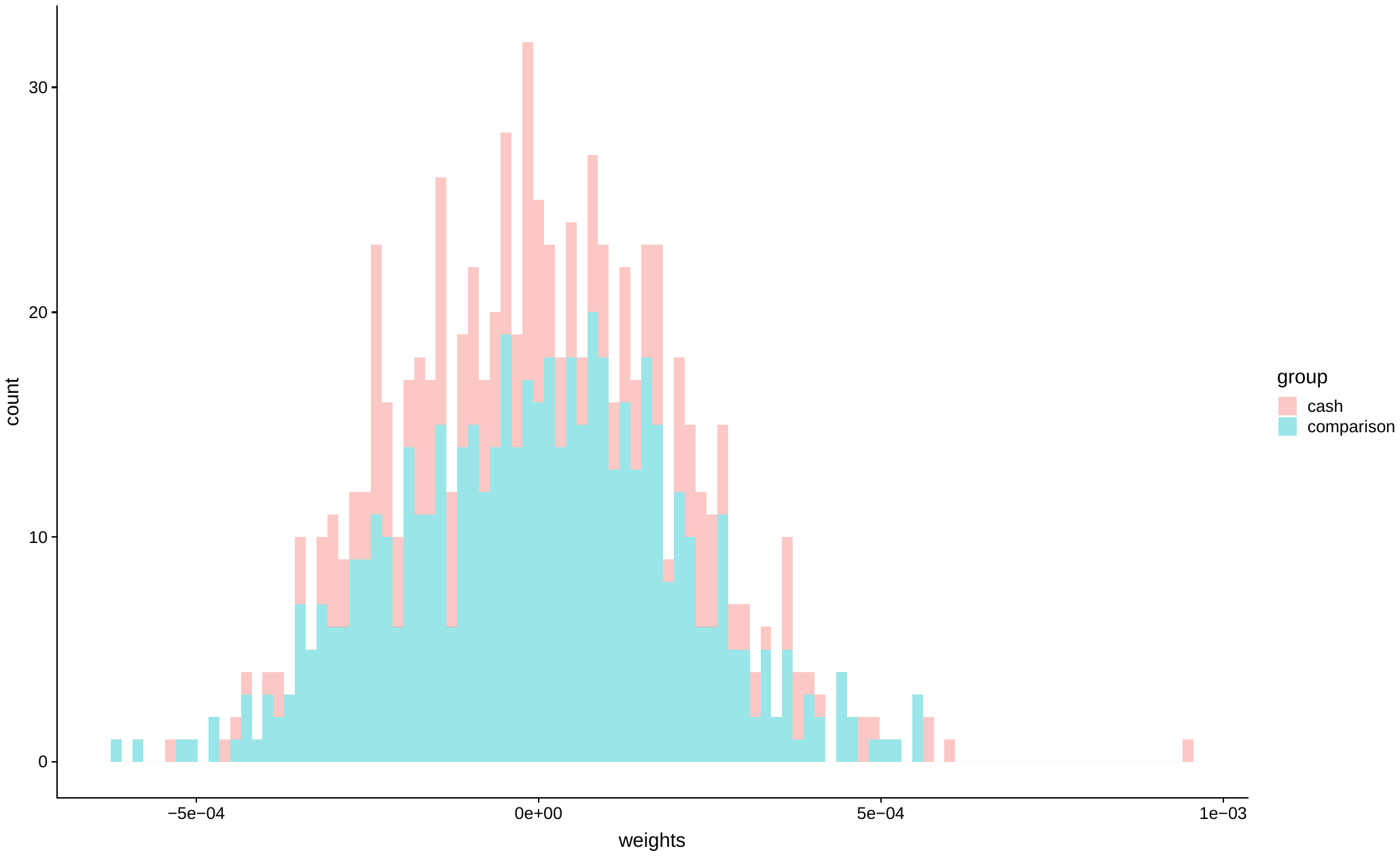

### Supplementary Figure 8. An illustration of the weights used to calculate two-way fixed effects estimates of the relationship between cash transfer programs and the proportion of people with HIV receiving antiretrovirals, with the weights being the residuals from a multivariable linear regression including country and year fixed effects, scaled by the sum of the squared residuals across all observations.

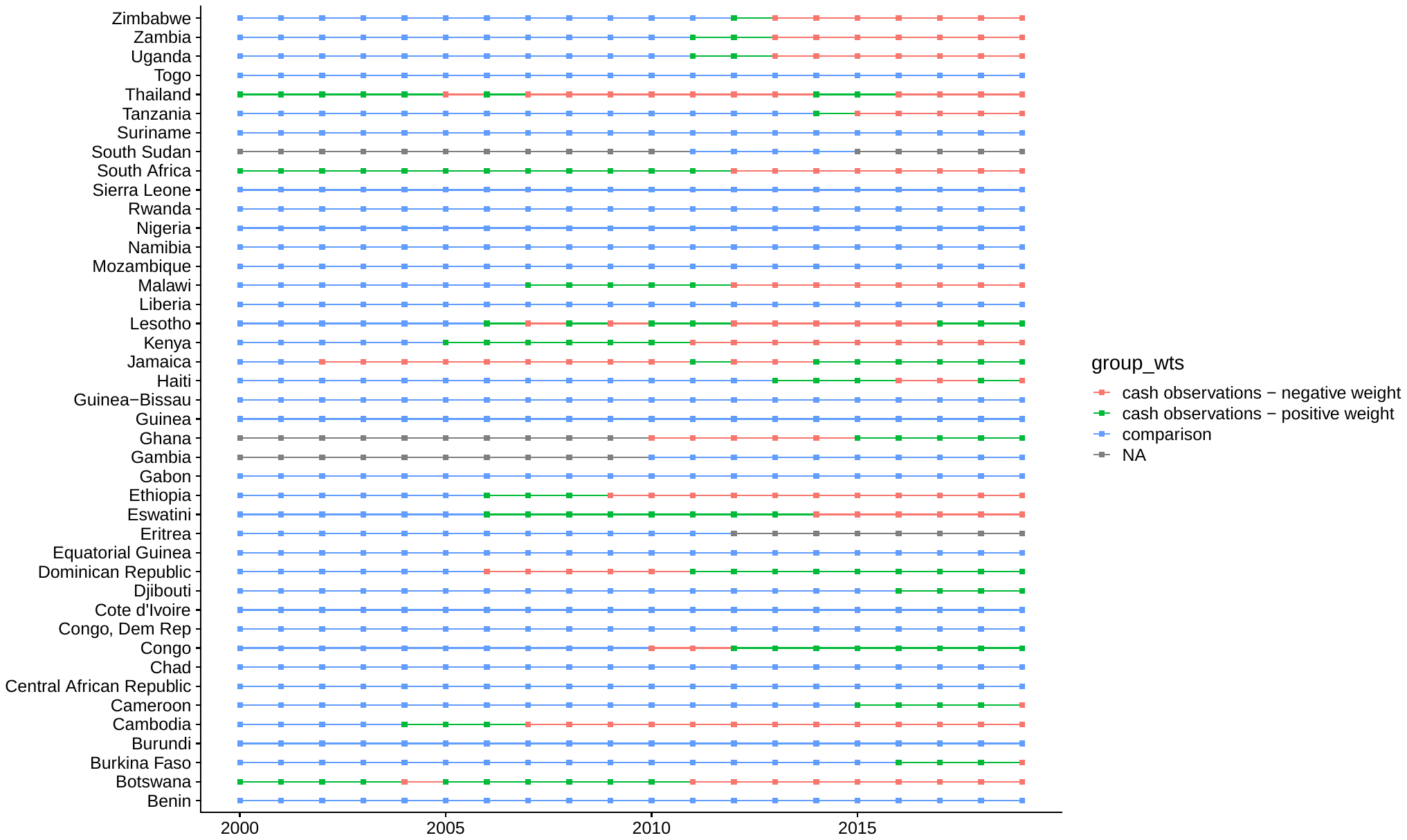

2.	United Nations Development Program Social Assistance in Africa Data Platform

. <https://social-assistance.africa.undp.org/country> (accessed 1/11/21.

3.	United Nations Economic Commission for Latin America and the Caribbean Non-contributory Social Protection Programs Database. <https://dds.cepal.org/bpsnc/cct> (accessed 1/11/21.

4.	The World Bank. ASPIRE: the Atlas of Social Protection Indicators of Resilience and Equity Program Inventory. <https://www.worldbank.org/en/data/datatopics/aspire> (accessed 1/11/21.

5.	Socialprotection.org Programme Database. <https://socialprotection.org/discover/programme> (accessed 1/11/21.

6.	United Nations Department of Economic and Social Affairs Population Division. Household Size and Composition. <https://population.un.org/Household/index.html#/countries/840> (accessed 1/13/21.

7.	The World Bank. World Development Indicators. <https://data.worldbank.org/data-catalog/world-development-indicators> (accessed 1/12/21.

8.	ICF International. Demographic and Health Survey Sampling and Household Listing Manual. MEASURE DHS, Calverton, Maryland, U.S.A.: ICF International, 2012.

9.	Goodman-Bacon A. Difference-in-differences with variation in treatment timing. *Journal of Econometrics* 2021.

10.	Callaway B, Sant’Anna PHC. Difference-in-Differences with multiple time periods. *Journal of Econometrics* 2020.

11.	Jakiela P. Simple Diagnostics for Two-Way Fixed Effects. *arXiv preprint arXiv:210313229* 2021.

12.	MacAuslan I, Farhat M, Bunly S, Craig R, Huy S, Singh P. Country-led Evaluation of the National Education Scholarship Programmes of the Ministry of Education, Youth and Sports in Cambodia (2015-2018) Final Report – Volume II: UNICEF, 2019.

13.	NOURISH Semi-Annual Progress Report: April 2016 - September 2016: Save the Children, 2016.

14.	Bank TW. Cameroon Social Safety Nets. <https://projects.worldbank.org/en/projects-operations/project-detail/P128534> (accessed 1/13/21.

15.	International Monetary Fund. Republic of Congo: Poverty Reduction Strategy Paper. IMG Country Report No. 12/242. Washington, D.C., 2012.

16.	Swaziland Old Age Grant Impact Assessment: Regional Hunger and Vulnerability Programme (RHVP), HelpAge International, UNICEF, 2010.

17.	Productive Safety Net Programme Phase IV Programme Implementation Manual. Addis Ababa: Ethiopia Ministry of Agriculture, 2014.

18.	Lesotho Social Assistance Budget Brief. Lesotho: Social Policy Section UNICEF, 2017.

19.	Uganda Launches Second Northern Uganda Social Action Project. 2010. <https://www.worldbank.org/en/news/feature/2010/02/09/uganda-launches-second-northern-uganda-social-action-project> (accessed 1/14/21.

20.	Office of the Prime Minister. UGANDA - SECOND NORTHERN UGANDA SOCIAL ACTION FUND (PRDP-NUSAF 2) - Environmental and Social Management Framework. Kampala, 2009.

21.	The World Bank. BI-Social Safety Nets (Merankabandi). <https://projects.worldbank.org/en/projects-operations/project-detail/P151835> (accessed 1/14/21.

22.	The World Bank. Chad Safety Nets Project. <https://projects.worldbank.org/en/projects-operations/project-detail/P156479?lang=en> (accessed 1/14/21.

23.	Bonilla J, Carson K, Kiggundu G, et al. Humanitarian Cash Transfers in the Democratic Republic of the Congo: Evidence from UNICEF’s ARCC II Programme Washington, D.C.: American Institutes for Research, 2017.

24.	Le Programme Filets Sociaux Productifs. <https://filetsociaux-ci.org/filets-sociaux/presentation/#:~:text=Le%20Programme%20Filets%20Sociaux%20Productifs,initial%20de%20la%20Banque%20Mondiale>. (accessed 1/14/21.

25.	Sederlof HSA. Implementation Completion Report (ICR) Review for GN Productive Social Safety Net Project: The World Bank, 2019.

26.	Rosas Raffo N. Sierra Leone - Sierra Leone Safety Nets Project : P143588 - Implementation Status Results Report : Sequence 03 (English): The World Bank, 2015.

27.	World Food Programme. WFP South Sudan Country Brief. 2017. <https://reliefweb.int/sites/reliefweb.int/files/resources/South%20Sudan%20Country%20Brief%20-%20%20January%202017%20OIM_0.pdf> (accessed 1/14/21.

28.	STIFLED IN DEVELOPMENT AND SCARED OF GETTING OLD. <https://www.socialwatch.org/node/11049> (accessed 1/14/21.
